# Maintaining excellence in care coordination during the COVID-19 pandemic and beyond: a survey of multidisciplinary healthcare teams in Ontario, Canada

**DOI:** 10.1101/2025.08.25.25334389

**Authors:** Donatus Mutasingwa, Joanne A. Permaul, Christopher Meaney, Jennifer Rayner, Stephen Marisette, Rahim Moineddin, Ross Upshur

## Abstract

**Introduction:** Patients with comorbidities have been shown to experience increased vulnerability during the COVID-19 pandemic, and to suffer disruption in their care coordination; having a multidisciplinary team is a care coordination strategy that can improve outcomes. The aim of this study was to describe the perspectives of team leads on coordination practices at their multidisciplinary health team (MHT) prior to, and during the COVID-19 pandemic.

**Methods:** Using a cross-sectional survey design, the Medical Home Care Coordination Survey for healthcare teams was distributed by email to executive directors or physician leads at all MHTs in Ontario, Canada. The outcome measures included the eight domains of care coordination, and participants’ rating of care coordination in general.

**Results:** The response rate was 58/241 (24%); 70% (95% CI: 0.50, 0.76) of teams reported using a validated method to identify complex patients in need of care coordination. High ratings for most items in the domains of care coordination prior to COVID-19 were maintained during the pandemic. Improvements can be made in providing patients with a copy of their care plan, making peer support accessible, and ensuring the timely inclusion of discharge summaries in the primary care record. Most participants (72%) rated care coordination in general at their MHT as *very good or excellent* prior to the pandemic; this decreased to 59% during the pandemic (p=0.016; 95% CI: −0.048, 0.31).

**Discussion:** To improve care coordination beyond the pandemic, providers should consider increasing the use of validated tools to identify patients with complex needs, and incorporating peer support systems to enhance care coordination efforts.

## Introduction

On March 11, 2020, COVID-19 was declared a global pandemic by the World Health Organization (WHO).^1^ By May 5, 2023 when the WHO announced an end to this public health emergency, there were more than 765 million confirmed cases, and over 6.9 million deaths worldwide.^2^ Initial studies have demonstrated that people with one or more comorbidities (e.g., diabetes, hypertension, etc.) were at a higher risk of COVID-related severe disease or mortality.^3,4^ One study showed that 88% of patients hospitalized for COVID-19 had one or more comorbidities.^5^ Beyond the increased vulnerability to COVID-19, patients with comorbidities were also likely to suffer from disruption in their care coordination.^5^ In order to reduce the risk of community COVID-19 transmission, primary care providers had to adjust the way that they provided services to their patients, for example through the adoption of virtual care or telemedicine in place of face-to-face visits. Care coordination still remained an essential function of primary care providers, and was even more important during the pandemic period, when these patients often needed coordination of other services (e.g., transportation, meals, etc.) in order for them to maintain social distance to reduce their COVID-19 risk.

Primary care is the most common setting within the Canadian healthcare system where patients with multiple chronic health conditions are managed, however these patients also require care from other health providers.^6^ With a decreasing number of healthcare providers, there is also the increased likelihood of healthcare fragmentation and subsequent adverse health outcomes and costs (e.g. increased health utilization, missed lab tests, medication errors and poor patient satisfaction). Care coordination has been identified as a promising tool for improving quality of care and reducing healthcare fragmentation.^7^

There is no consensus on how to define care coordination however, the U.S. Agency for Healthcare Research and Quality provides a widely used definition: *“the deliberate organization of patient care activities between two or more participants (including the patient) involved in a patient’s care to facilitate the appropriate delivery of health care services. Organizing care involves the marshalling of personnel and other resources needed to carry out all required patient care activities, and is often managed by the exchange of information among participants responsible for different aspects of care”*.^8(p.41)^

To better manage patients with complex health needs, such as those with multiple comorbidities, identifying patients in greatest need of coordinated care has been determined to be a top priority.^9^ Prior to the COVID-19 pandemic, studies estimated that up to 40% of patients with chronic and multiple comorbidities in Canada have experienced one or more care coordination gaps.^9^ According to a 2015 Commonwealth Fund Survey, Canada ranked below average in terms of care coordination for patients with complex needs, with only about 60% receiving the required coordination compared to an average of 70% in other countries.^10^ A more recent Commonwealth Fund survey showed that although 60% of primary care physicians in Canada screened their patients for social need; only 43% coordinated with social services.^11^ Previous studies have identified several barriers that affected the quality of care coordination including poor communication between primary care providers and specialists, and lack of personnel.^11,12^

Care coordination has been associated with better overall patient satisfaction with chronic disease care and can potentially reduce healthcare costs.^13,14^ With the projected increase in people with multiple chronic diseases, there is a greater need to continually improve how care coordination is practiced in order to better optimize care for this group of vulnerable patients, and thereby contain or reduce healthcare costs. The need was even more urgent given the service disruption caused by the COVID-19 pandemic.

Having a multidisciplinary team has been shown to be one of the care coordination strategies that can improve outcomes (e.g. service continuity, hospitalization, and mortality) among a select group of patients,^7^ and more recently found to improve functional difficulties among patients with multimorbidity.^15^ In the province of Ontario, Canada, Family Health Teams (FHTs) and Community Health Centres (CHCs) are examples of such multidisciplinary healthcare teams (MHTs). Although these FHTs and CHCs have funding to handle some of the increased demand for care coordination, there are few studies that have examined the current practice within such organizations. A study by Carroll et al., for example, showed that 20% – 60% of patients seen at academic FHTs ranked various components of care coordination below the minimum expected level.^16^ This study, however, was limited to academic FHTs and did not examine the existing care coordination practices among the surveyed FHTs. Despite increasing interest in care coordination amongst policy makers and researchers, there is still no consensus on how to run an efficient and effective care coordination program.^7^

Few studies have addressed the effects of the COVID-19 pandemic on care coordination. A recent study by Gaucher et al. surveyed a convenience sample of independent midwives in France to determine the effects of the initial COVID-19 lockdown on care coordination compared to prior to the pandemic.^17^ The study results showed that 65% of midwives experienced new difficulties with patient referrals during the pandemic, particularly to social workers and specialist physicians, and 71% encountered new difficulties collaborating with hospitals, on issues such as health data transmission, organizing unscheduled care, and the implementation of protocols.^17^ Recently, Whitebird et al. conducted a qualitative study with patients receiving care coordination to determine if their healthcare, social and financial needs were affected by the COVID-19 pandemic. Analysis of data from semi-structured interviews with 19 patients showed that although patients were feeling disconnected from their family, peers, and community, care coordinators were able to assist patients in accessing resources so that they could maintain their overall health and wellbeing.^18^

### Objective

The objectives of this study were to describe Ontario MHT leads’ perspectives on existing care coordination practices for patients with multiple comorbidities, and to determine if these practices were impacted during the COVID-19 pandemic.

### Relevance

Care coordination has been associated with better overall patient satisfaction with chronic disease care and can potentially reduce healthcare costs.^14^ With the projected increase in people with multiple chronic diseases, there is a greater need to continually improve how care coordination is practiced in order to better optimize care for this group of vulnerable patients, and thereby contain or reduce healthcare costs. This study is relevant because it provides an overview of the care coordination perspective of providers in Ontario. The study findings will provide the baseline data required to design an intervention study geared towards improving care coordination practices in MHTs, and will provide empirical evidence for best practice recommendations with respect to care coordination in future public health emergencies.

## Methods

Ontario MHTs (N=241) were invited to participate in this survey, from October 2020 to February 2021. Family Health Team executive directors or physician leads with publicly available email addresses (N=136), as well as executive directors of Community Health Centres (CHC) listed with the Alliance for Healthier Communities (N=105), were contacted to complete a survey on their perspectives of care coordination practices at their MHT. The CHC organizations included community FHTs, community health centres, and nurse practitioner (NP) led clinics. Email addresses for FHT executive directors were obtained through internet searches and invitations were sent out by the study coordinator (JP) on behalf of the Principal Investigator (DM). Invitations to CHCs were emailed by the Director of Research and Evaluation at the Alliance (JR).

### Survey Instrument

The 32-item Medical Home Care Coordination Survey for healthcare teams (MHCCS-H) assesses elements of care coordination from the healthcare team perspective, and has been found to be valid and reliable.^20^ This tool addresses the eight domains of care coordination which includes accountability, IT capacity, plan of care, follow-up plan of care, self-management, communication, link to community resources and care transitions (Box 1).^19^ We modified the MHCCS-H questions to ask respondents about their care coordination practices prior to and during the COVID-19 pandemic (Appendix A). Each item was rated on a 5-point Likert scale ranging from *strongly disagree* to *strongly agree, never* to *always*, or *poor* to *excellent*. Supplementary questions were also developed to determine practice characteristics, and the tools most commonly used by MHTs to identify patients in need of enhanced care coordination. The MHCCS-H and supplementary questions were converted to an online survey using the Qualtrics_®_ platform; a hyperlink to the survey was included in the invitation email message.

**Box 1.**
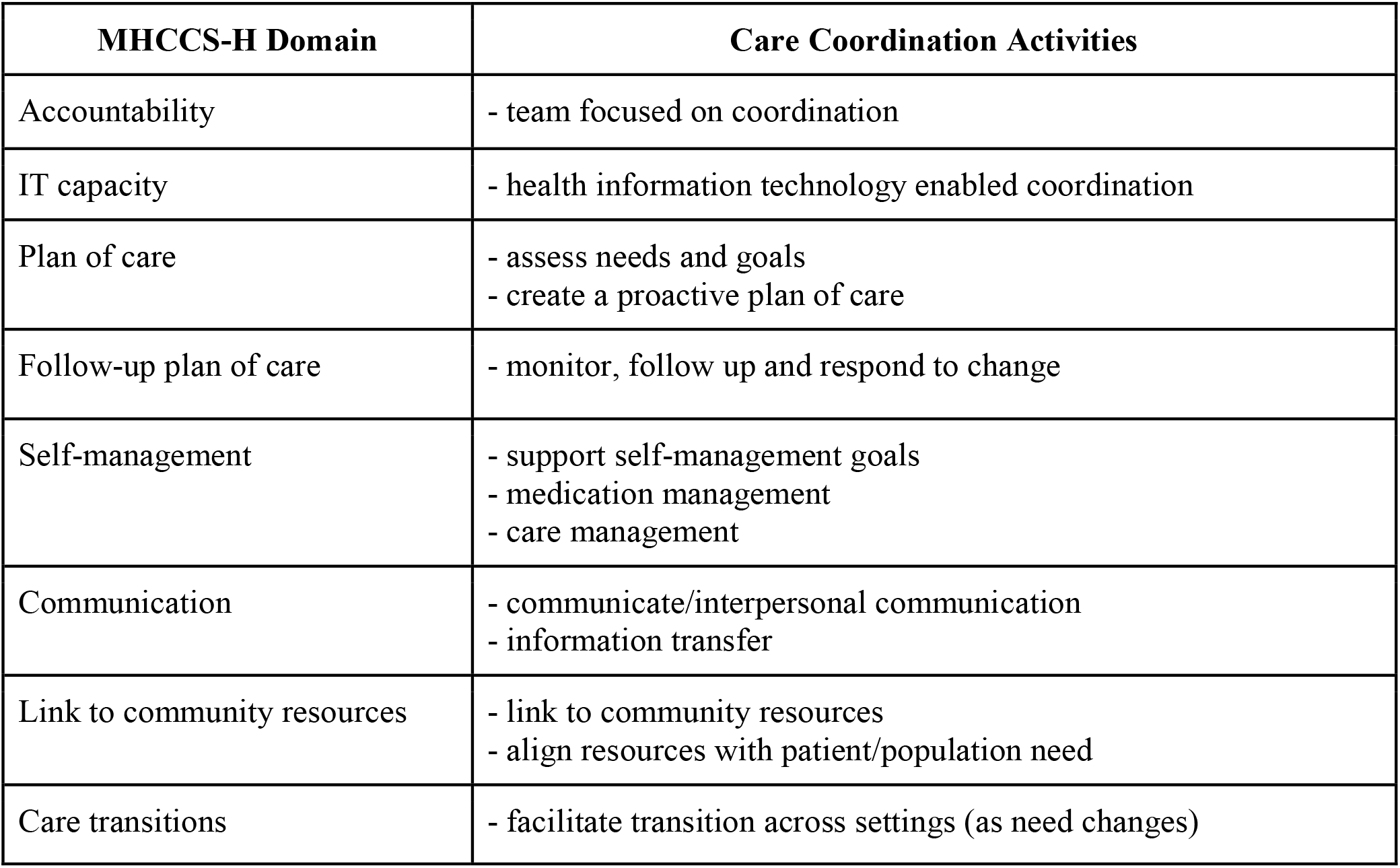
The 8 Domains of Care Coordination and Corresponding Activities^19^.

| MHCCS-H Domain | Care Coordination Activities |
| --- | --- |
| Accountability | - team focused on coordination |
| IT capacity | - health information technology enabled coordination |
| Plan of care | - assess needs and goals<br>- create a proactive plan of care |
| Follow-up plan of care | - monitor, follow up and respond to change |
| Self-management | - support self-management goals<br>- medication management<br>- care management |
| Communication | - communicate/interpersonal communication<br>- information transfer |
| Link to community resources | - link to community resources<br>- align resources with patient/population need |
| Care transitions | - facilitate transition across settings (as need changes) |

### Data Analysis

The survey data were analyzed using IBM SPSS Version 27. Counts, proportions/percentages and medians were used to analyze the data descriptively, and McNemar’s tests compared responses prior to, and during the COVID-19 pandemic. For items rated on a 5-point Likert scale, the top two response were combined (e.g., *somewhat agree/agree, usually/always, very good/excellent*) and compared to all other responses. The study outcomes included items under the 8 domains of care coordination, and an overall rating of care coordination.

### Research Ethics

Ethics approval for this study was received from the Oak Valley Health Research Ethics Board in Ontario, Canada (REB #119-2009).

## Results

A total of 58/241 (24%) executive directors or physician leads at Ontario MHTs completed the study; 53 (91%) of these respondents provided information about their organization (Table 1). Most of the organizations represented in the data were CHCs (58%); FHTs accounted for 32%, while 8% were Community (governed) FHTs and 2% were nurse practitioner led clinics. Only 16% of MHTs were academic sites, and while 98% of respondents reported that they had an electronic medical record (EMR) at their site, only 79% used it to identify patients at high risk or those in need of care coordination.

**Table 1.** Practice Characteristics by Type of Organization (N=53)

| Characteristic | Organization Type* |  |  |  | Total<br>(N=53) |
| --- | --- | --- | --- | --- | --- |
|  | FHT<br>(N=17) | Community FHT<br>(N=4) | CHC<br>(N=31) | NP Led Clinic<br>(N=1) |  |
| Length of Operation (Median # Years) | 12.5 | 13.0 | 25.0 | 9.0 | 13.5 |
| Median # Satellite FHT Sites | 3.0 | 2.0 | 2.5 | - | 3.0 |
| Median # Rostered Patients | 19,000 | 11,750 | 5,000 | 2,550 | 6,000 |
| Median # Family Doctors or Nurse Practitioners | 14.0 | 12.0 | 9.0 | 4.0 | 9.0 |
| Median # Allied Health Professionals | 12.0 | 7.25 | 12.0 | 6.0 | 12.0 |
|  | # (%) | # (%) | # (%) | # (%) |  |
| Academic Site: |  |  |  |  |  |
| Yes | 6 (40.0) | 0 | 2 (6.7) | 0 | 8 (16.0) |
| No | 9 (60.0) |  | 28 (93.3) |  | 42 (84.0) |
| Primary Person Involved with Care<br>Coordination / Case Management: |  |  |  |  |  |
| Physician | 4 (23.5) | 2 (50.0) | 6 (20.0) | 0 | 12 (23.1) |
| Case Manager | 0 | 0 | 4 (13.3) | 0 | 4 (7.7) |
| Nurse | 7 (41.2) | 1 (25.0) | 7 (23.3) | 1 (100.0) | 16 (30.8) |
| Other | 4 (23.5) | 0 | 12 (40.0) | 0 | 16 (30.8) |
| Unsure | 2 (11.8) | 1 (25.0) | 1 (3.3) | 0 | 4 (7.7) |
| Have an EMR? |  |  |  |  |  |
| Yes | 16 (94.1) | 4 (100.0) | 31 (100.0) | 1 (100.0) | 52 (98.1) |
| No | 1 (5.9) | 0 | 0 | 0 | 1 (1.9) |
| Using EMR to Identify Patients Who Need Care<br>Coordination / High Risk Pts: |  |  |  |  |  |
| Yes | 16 (94.1) | 4 (100.0) | 20 (66.7) | 1 (100.0) | 41 (78.8) |
| No | 1 (5.9) | 0 | 10 (33.3) | 0 | 11 (21.2) |
| Using at Least One Care Coordination Tool: |  |  |  |  |  |
| Yes | 13 (76.5) | 2 (50.0) | 21 (67.7) | 1 (100.0) | 37 (69.8) |
| No | 4 (23.5) | 2 (50.0) | 10 (32.3) | 0 | 16 (30.2) |

\*Sample sizes may vary due to missing data.

Most teams (77%) had non-physician practitioners responsible for care coordination. The proportion of teams that reported using electronic data to identify patients with complex health needs was 88% pre-pandemic and 81% during the pandemic. Despite the widespread use of electronic data, only 70% of the teams reported using at least one validated method to identify complex patients in need of care coordination. Counting the total number of comorbidities and medications was the most reported method used to identify patients in need of care coordination and was used by 64% of teams.

Table 2 displays the frequency distributions from the eight domains of care coordination in Ontario MHTs before and during COVID-19, and the results from the McNemar’s tests comparing dependent proportions estimated prior to, and during the pandemic. Overall, the results show that the high ratings of the domains of care coordination prior to COVID-19 were maintained during the pandemic. There was only one significant change – the majority of team leads (72%) rated the care coordination at their MHT as *very good* or *excellent* prior to the pandemic, but this proportion dropped to 59% during the pandemic (p=0.016). This is an interesting finding because although there was no significant change in ratings in any of the eight domains of care coordination during COVID-19 compared to prior to the pandemic, there was a significant decrease in the proportion of team leads who highly rated care coordination in general at their MHT during the pandemic.

**Table 2.**
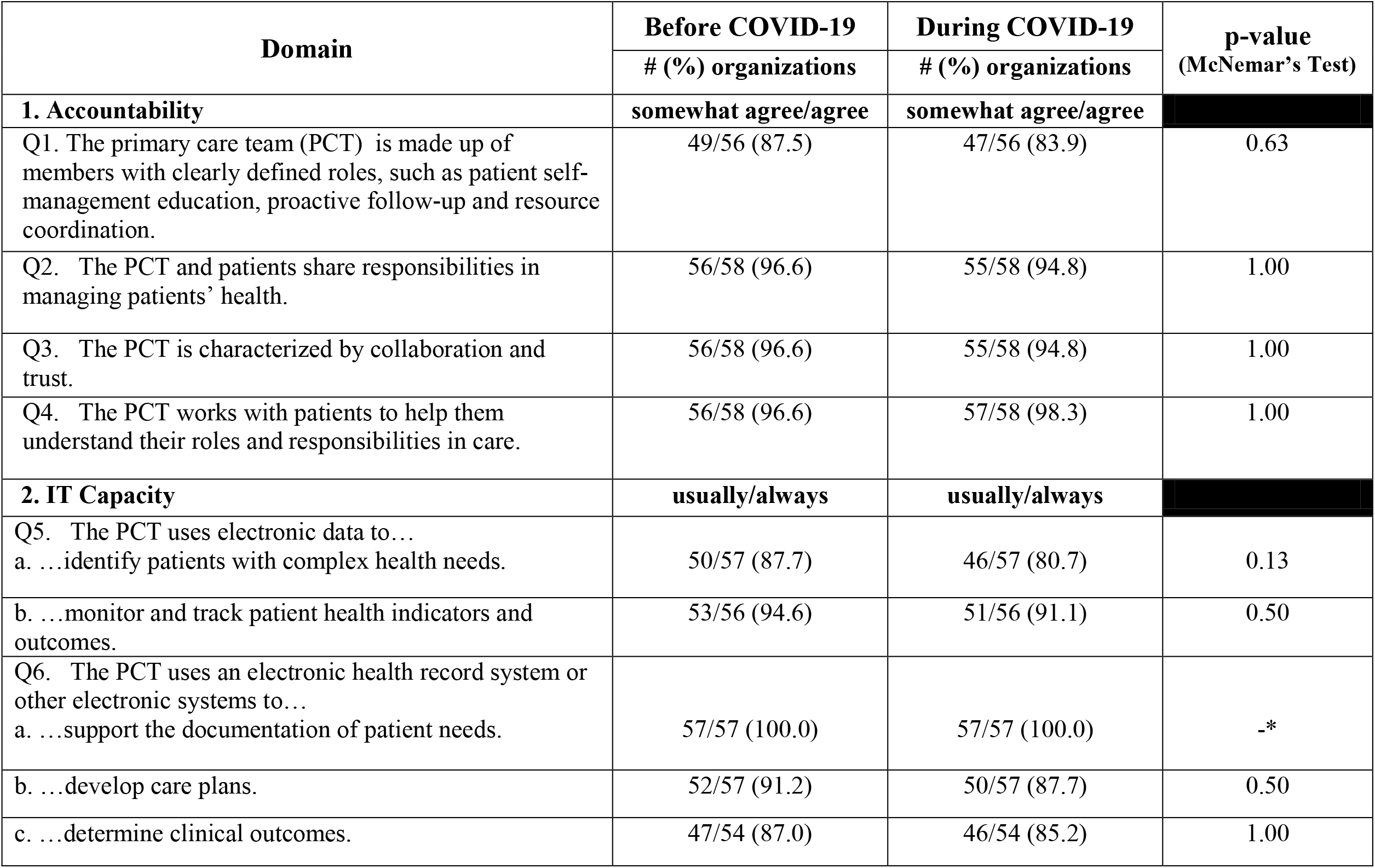
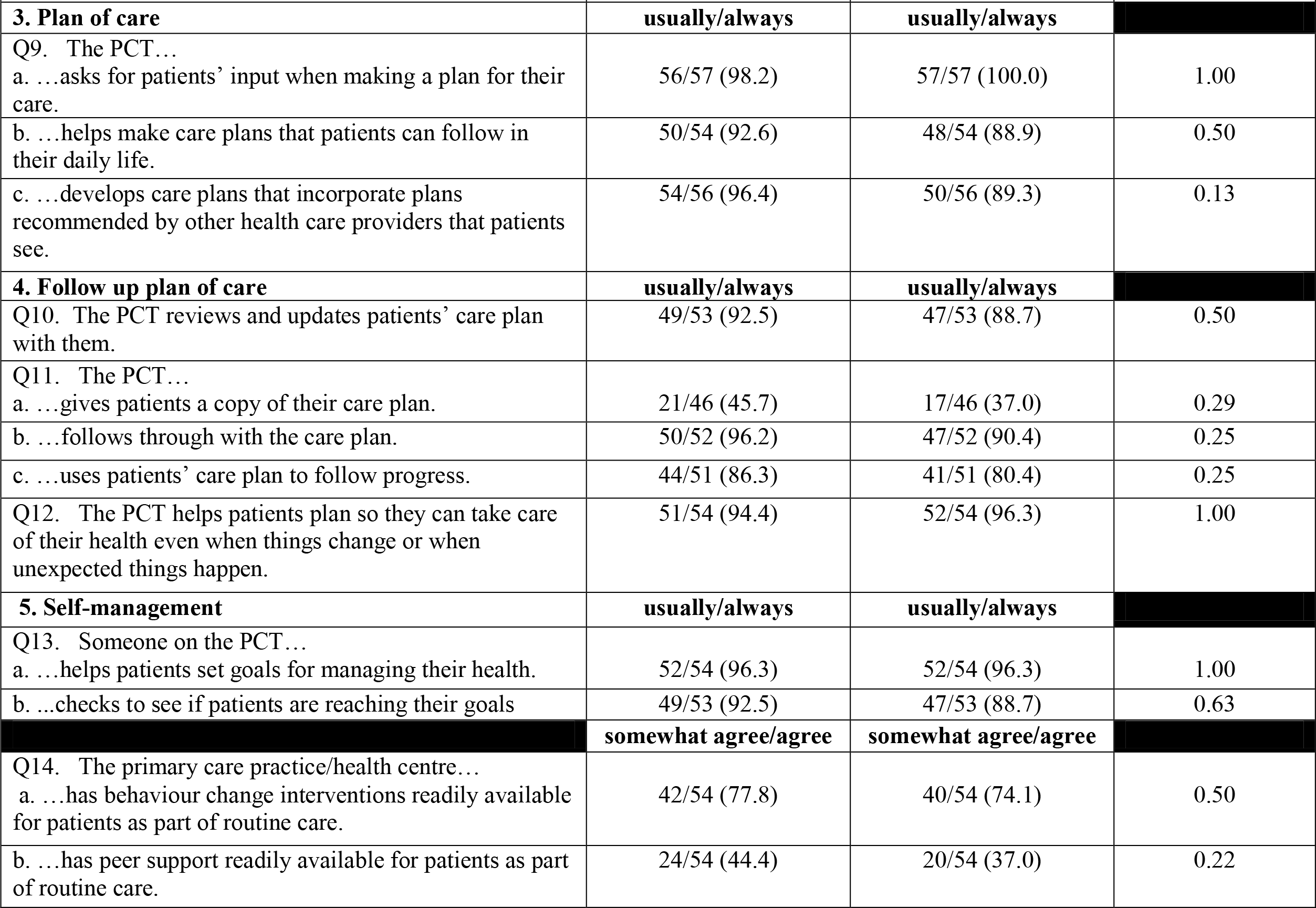
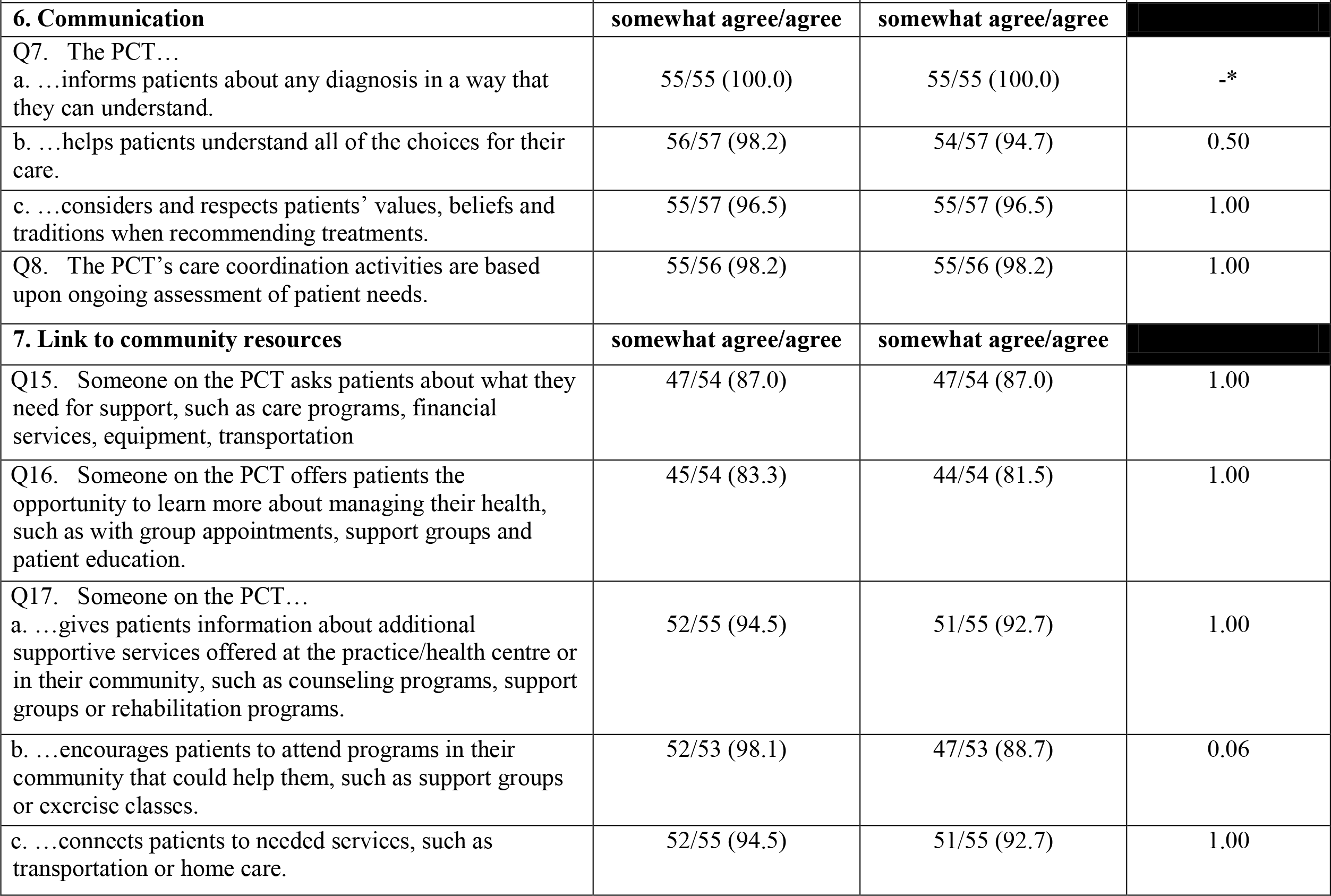
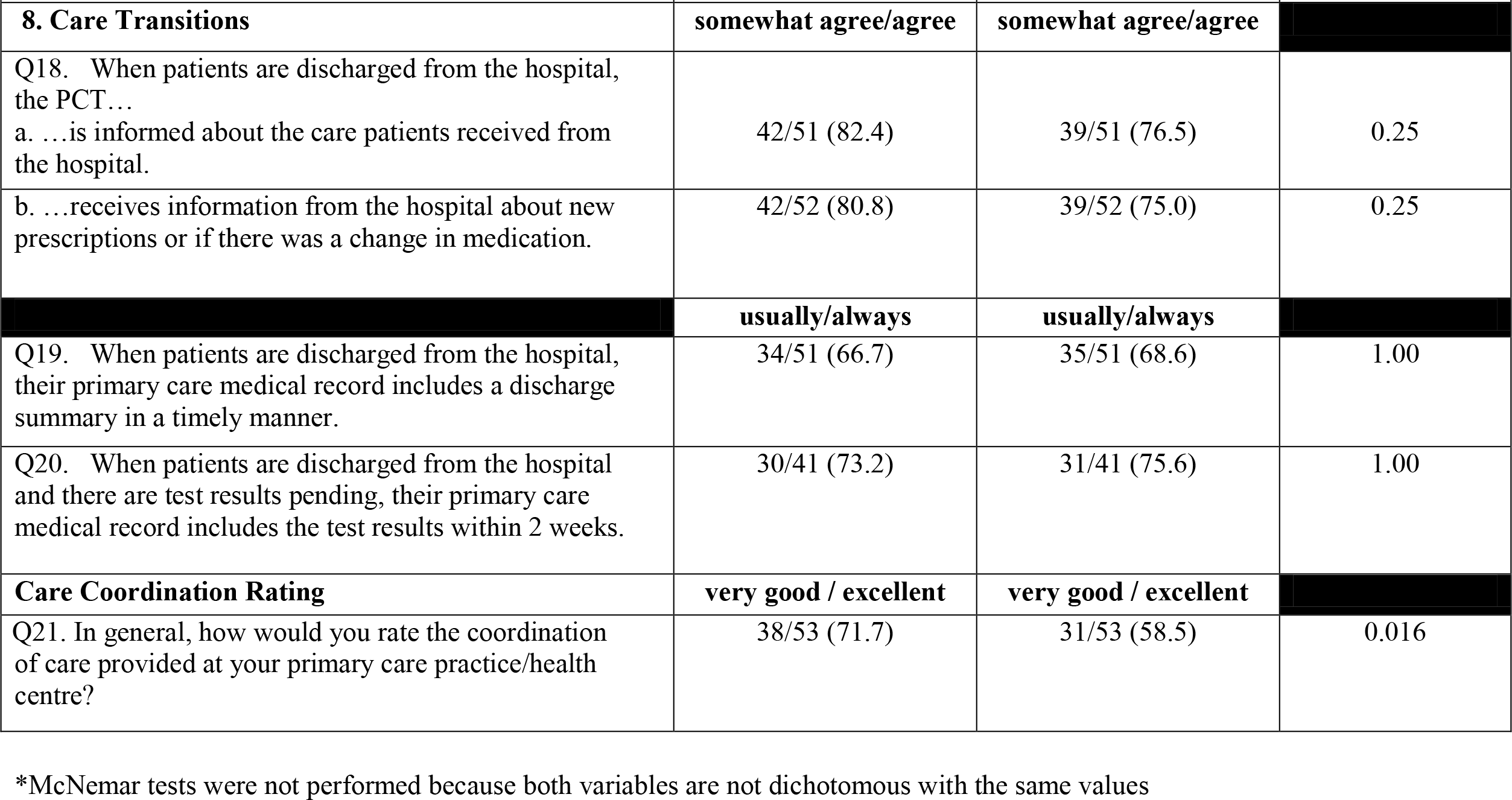
The Eight Domains of Care Coordination in Ontario Multidisciplinary Healthcare Teams: Before and During COVID-19 (N=58)

## Discussion

When the COVID-19 pandemic was declared in March 2020, primary care providers were required to adjust how they provided services in order to reduce the risk of community transmission. Multidisciplinary health care teams in Ontario, Canada were challenged to maintain excellence in the care coordination services provided during the pandemic while minimizing the risk of COVID-19 transmission.

This cross-sectional survey examined MHT leaders’ perspectives on care coordination prior to and during the COVID-19 pandemic, using a modified 32-item MHCCS-H instrument. The results showed that none of the items that were rated under the eight domains of care coordination had changed significantly during COVID-19, compared to prior to the pandemic.

However, when asked in a separate question to rate the care coordination provided at their MHT in general, there was a significant decrease in the percentage of those who highly rated care coordination (as *excellent* or *very good*) during the pandemic, compared to before the pandemic.

The accountability domain of the MHCCS-H reflects the collaboration between members of the health care team and the patient, and their ability to share responsibilities to manage the patient’s health.^19^ The ratings of items in this domain were very high prior to COVID-19, and remained high during the pandemic, indicating that teams were able to maintain connections with their patients. There is room for improvement however, in clearly defining the role of each team member, particularly during a public health emergency.

All of the MHTs were using an EMR or other electronic system to document patient needs prior to and during the pandemic, and most used electronic data to monitor the health indicators and outcomes of their patients, however not all MHTs were using the EMR to identify patients with complex healthcare needs. It is important to identify patients with complex needs early in order to offer interventions that may improve their quality of life, and to minimize unnecessary use of healthcare services and related costs.^20^ A number of tools have been developed to identify patients with complex health needs who may require additional services in a variety of contexts, which could be used to predict hospitalization, higher utilization of healthcare services, and associated costs.^20^

Almost all of the MHTs included patients’ input in the development of a plan that can be easily followed in their daily lives – this did not change during the COVID-19 pandemic. Fewer than half of the MHTs however, gave patients a copy of their care plan prior to the pandemic and while this number decreased further during the pandemic, most MHTs reported that they usually or always followed through with the care plan and used the plan to follow their patients’ progress.

Studies have demonstrated that peer support can have a positive impact on the self-management of chronic diseases.^21–22^ Individuals with the same or similar condition, who provide support by sharing experiences, knowledge and advice, and giving emotional support, can be a source of encouragement for patients with chronic illnesses.^21–22^ The results of the current study show that fewer than half of the respondents *agreed* or *somewhat agreed* that their MHT had peer support readily available for patients as part of their routine care prior to COVID-19, and this number dropped further during the pandemic. A recent scoping review showed that online peer support programs can provide the same impact as face-to-face interventions, with significant improvements in social participation, empowerment, and health-related behaviours.^21^ This method of peer support should be considered, particularly during a public health emergency, to increase accessibility and potentially decrease the stigma associated with face-to-face interactions.

When it came to connecting patients with community resources, the MHT ratings were high prior to and during the COVID-19 pandemic. However, there is room for improvement in asking patients about their needs for support, and providing them with opportunities to learn about managing their health. Although not significant, there was a 10% decline during the pandemic in the proportion of MHTs that encouraged patients to attend community programs that could assist them. This finding is not surprising given that many programs such as exercise classes and support groups were suspended during the pandemic. The data on care transitions showed that the majority of MHTs are informed when patients are discharged from the hospital and that they receive information about their care and medication adjustments. Discharge summaries and test results however, could be included in the medical record in a timelier manner.

Although the items in the eight domains of care coordination did not change significantly during the pandemic, respondents’ overall ratings of care coordination at their MHT dropped significantly compared to prior to the pandemic. According to the Ontario COVID-19 Science Advisory Table, delays and deferrals of preventive and ongoing care during the pandemic made it more difficult to provide care to patients with increasingly complex care needs, adding stress to the already strained healthcare system, and contributing to the erosion of the quality of primary care in Ontario.^23^ As a result, respondents’ perceptions of the care provided at their MHT may have been impacted by their impressions of the overall state of health care in Ontario, and the ongoing issues that were magnified during the pandemic.

### Limitations

This study had some limitations. The low response rate could be related to the online distribution of the survey as this method typically yields lower response rates that other modes of survey distribution,^24^ or it could be a result of survey fatigue due to the increase in surveys conducted during COVID-19.^25^ It is also possible that incorrect email addresses were extracted from the web searches, or there was a general lack of interest in care coordination from those surveyed. Given the small sample size, there was not sufficient power to conduct multivariate analyses to examine if any of the practice characteristics were associated with ratings on the MHCCS-H. For many of the domain items, the ratings were already very high prior to COVID-19, creating a *ceiling effect* and making it difficult to detect any changes during the pandemic. In addition, the study design, which required respondents to rate each item prior to and during the pandemic at a single time point, may have led to recall bias and inaccurate responses. A longitudinal study that asked for ratings on care coordination at distinct time points would have been a stronger study design.

Although multidisciplinary health teams in Ontario maintained high ratings on most of the items in the eight domains of care coordination during the COVID-19 pandemic, we recommend that to further improve care coordination beyond the pandemic, healthcare providers should consider increasing the use of validated tools to identify patients with complex needs such as the Lace Index Scoring Tool, the Predictive Repetitive Admission (PRA) Tool and others, as well as providing accessibility to peer support systems to enhance care coordination efforts. Future studies should examine in-depth characterization of care coordination practices and how various teams are using their resources to obtain optimal outcomes for patients In addition, more work is required to evaluate the effectiveness of existing care coordination tools and interventions.

## Data Availability

All data produced in the present work are contained in the manuscript.

## Appendix A: Modified MHCCS-H and Supplementary Questions

### The Medical Home Care Coordination Survey (MHCCS-H)^1^

#### SURVEY INSTRUCTIONS

- Do your best to answer each question based on your opinion of the care coordination provided at your primary care practice/health centre in the **last 6 months**
- Please **only** select **one answer per question** unless specified otherwise
- Please answer all questions by selecting the box to the left of your answer
- Please answer all questions honestly and completely

**Please read this to better understand the survey questions:**

**CARE COORDINATION**^**2**^ is the “deliberate organization of patient care activities between two or more participants (including the patient) involved in a patient’s care to facilitate the appropriate delivery of health care services. Organizing care involves the marshaling of personnel and other resources needed to carry out all required patient care activities and is often managed by the exchange of information among participants responsible for different aspects of care”.

^*^ References:

1. Zlateva I, Anderson D, Coman E, et al. Development and validation of the Medical Home Care Coordination Survey for assessing care coordination in the primary care setting from the patient and provider perspectives. *BMC Health Serv Res* 2015; 15: 226.

2. Macdonald KM, Sundaram V, Bravata DM, et al. Closing the quality gap: a critical analysis of quality improvement strategies (Vol. 7 Care Coordination). Rockville (MD): Agency for Healthcare Research and Quality (US); 2007 Jun. (Technical Reviews, No. 9.7.) 3, Definitions of Care Coordination and Related Terms. Available from: https://www.ncbi.nlm.nih.gov/books/NBK44012/

#### PLEASE BEGIN THE SURVEY

1 **The primary care team is made up of members with clearly defined roles, such as patient self management education, proactive follow up and resource coordination**.

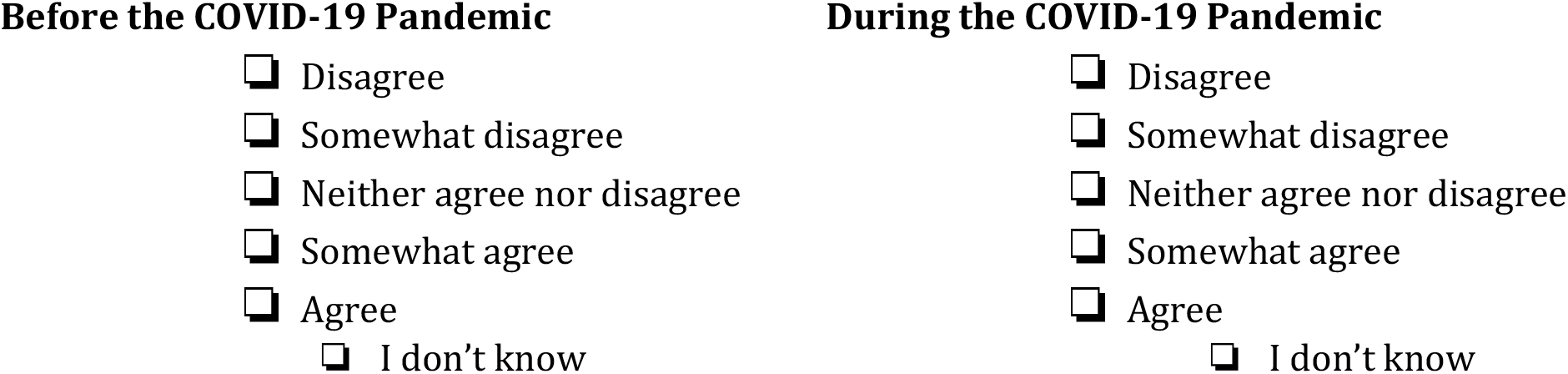

2 **The primary care team and patients share responsibilities in managing patients’ health**.

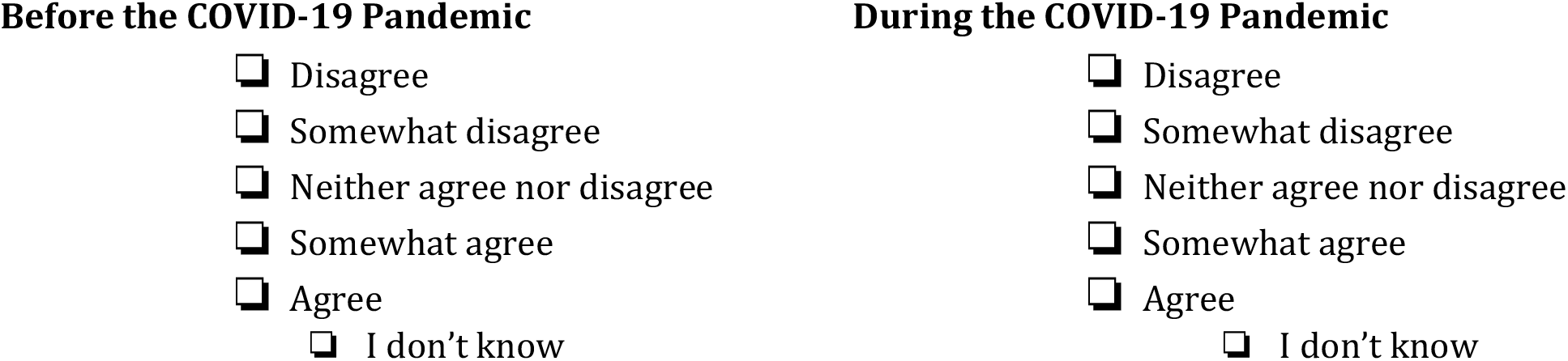

3 **The primary care team is characterized by collaboration and trust**.

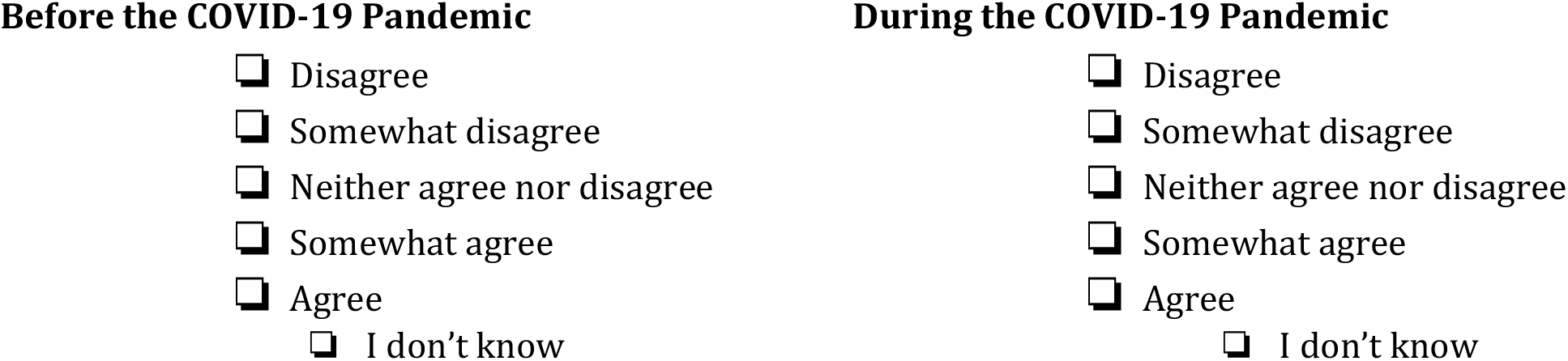

4 **The primary care team works with patients to help them understand their roles and responsibilities in care**.

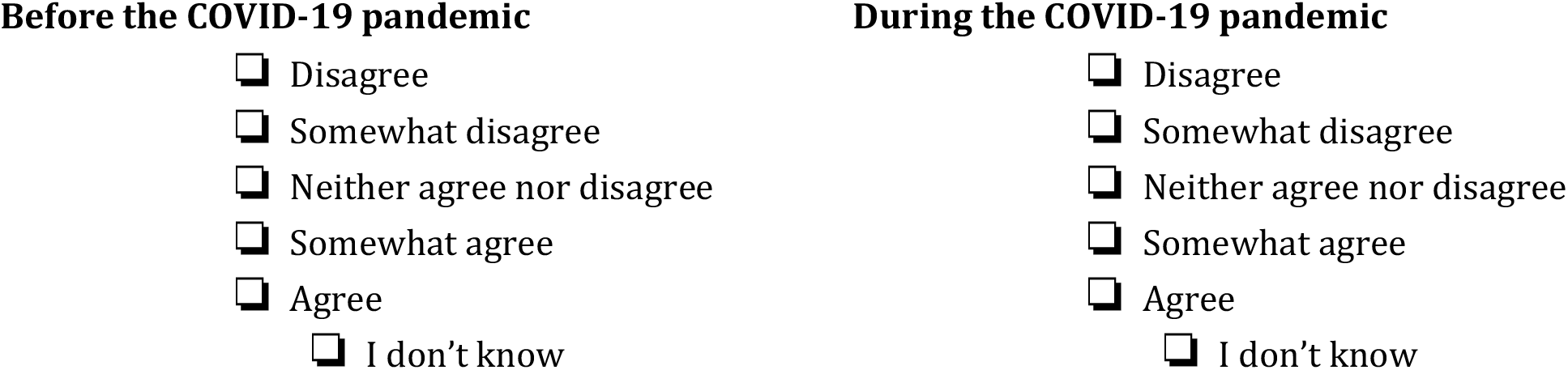

5 **The primary care team uses electronic data to…**

a **…identify patients with complex health needs**.

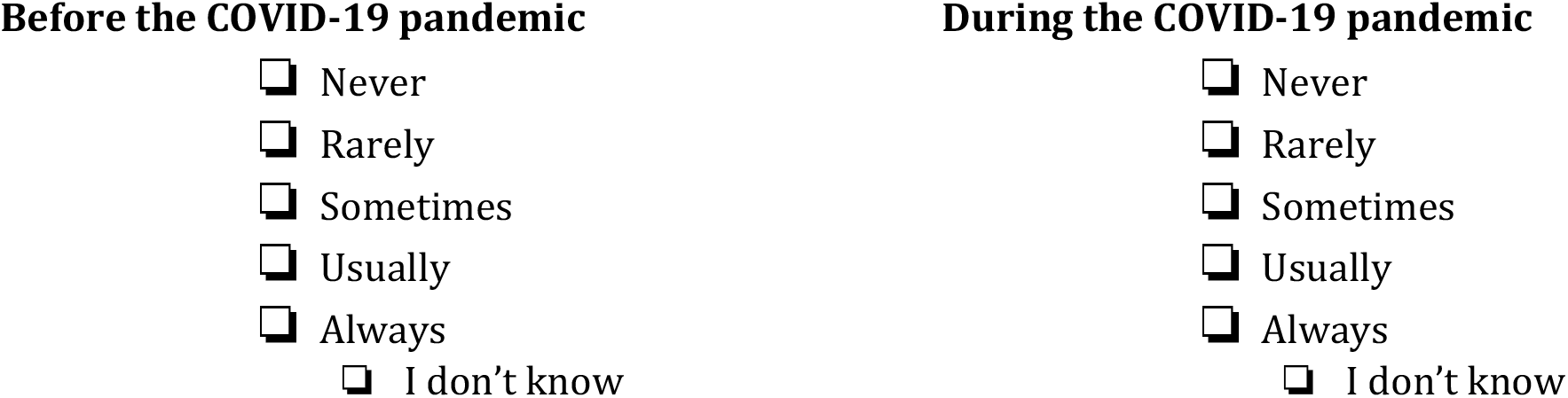

b **…monitor and track patient health indicators and outcomes**.

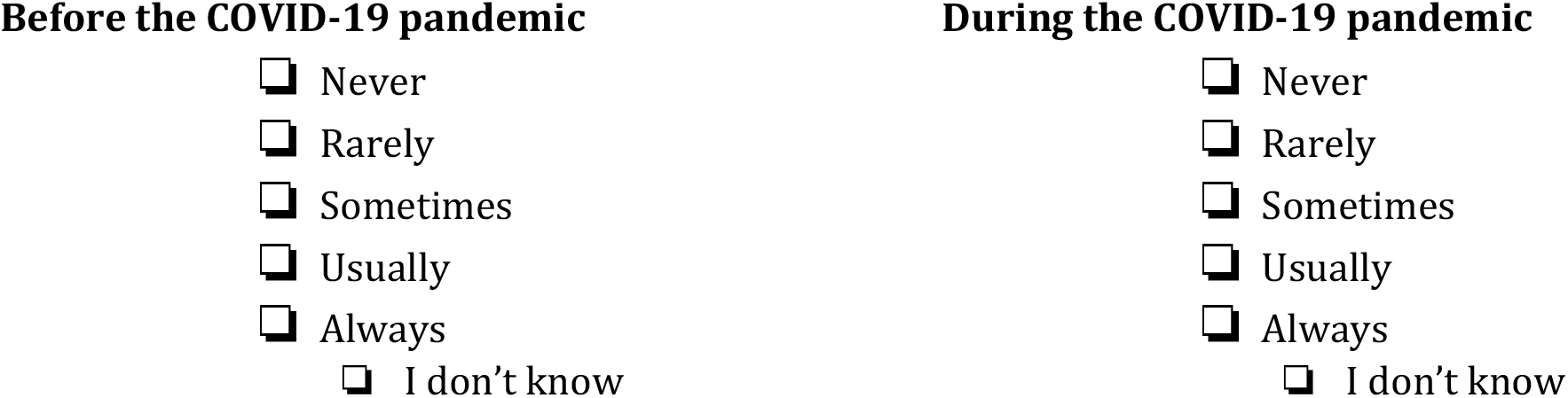

6 **The primary care team uses an electronic health record system or other electronic systems to…**

a **…support the documentation of patient needs**.

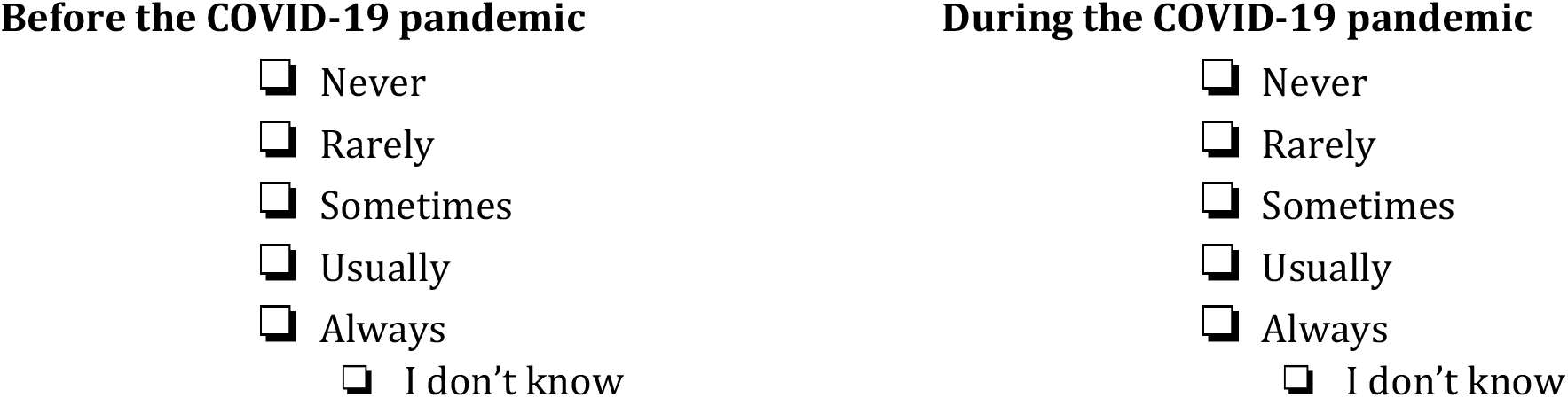

b **…develop care plans**.

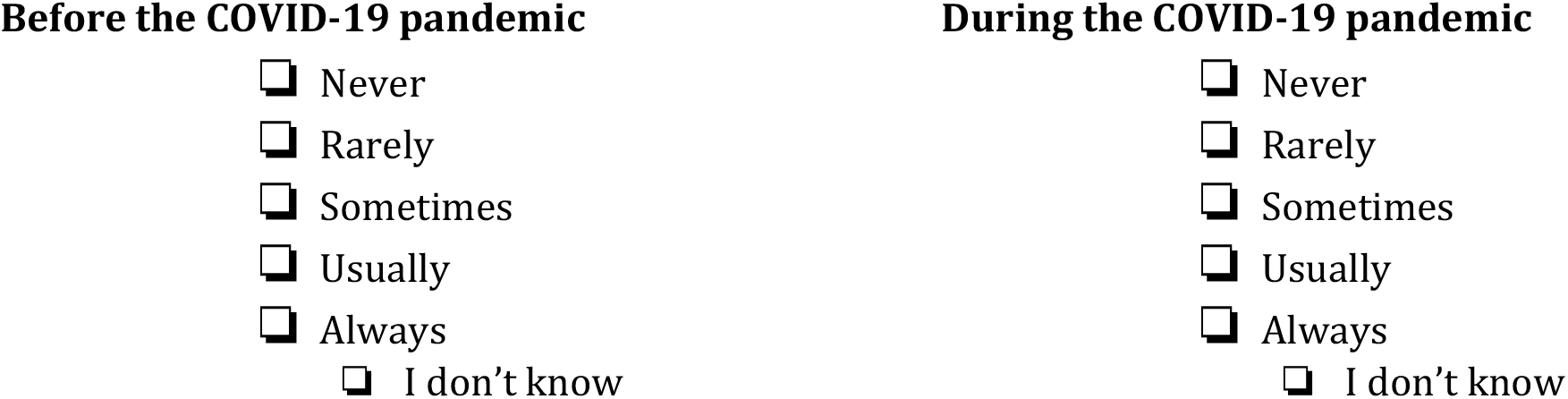

c **…determine clinical outcomes**.

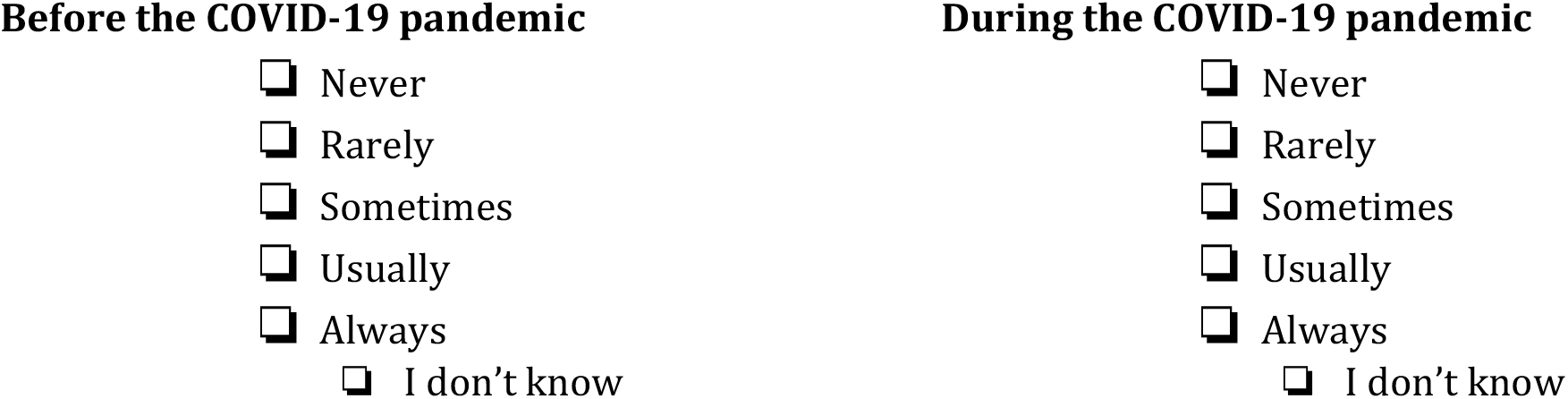

7 **The primary care team…**

a **…informs patients about any diagnosis in a way that they can understand**.

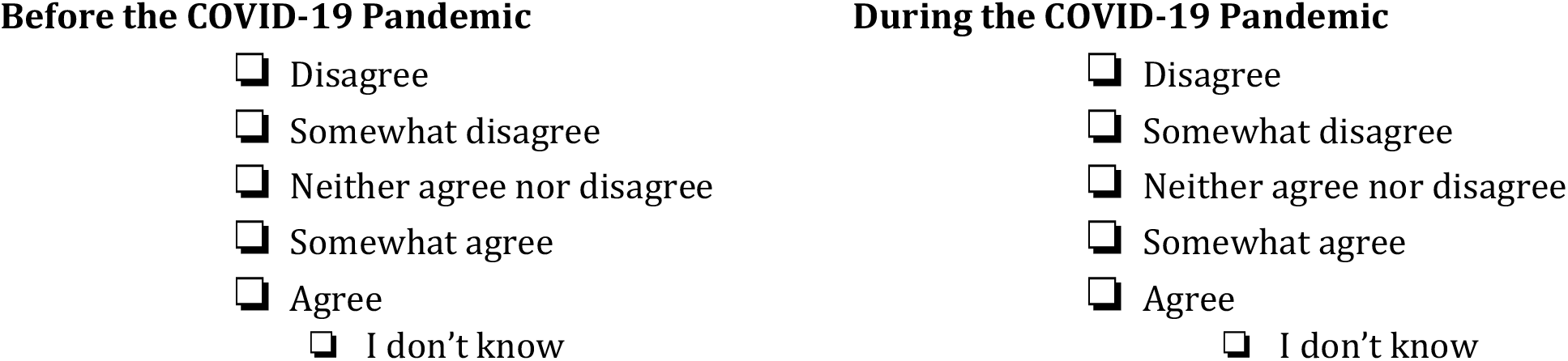

b **…helps patients understand all of the choices for their care**.

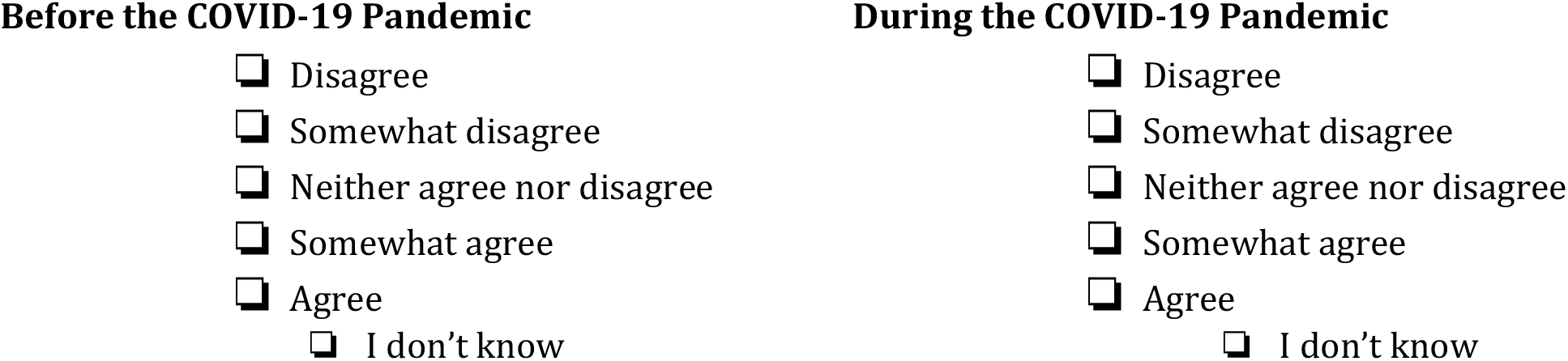

c **…considers and respects patients’ values, beliefs and traditions when recommending treatments**.

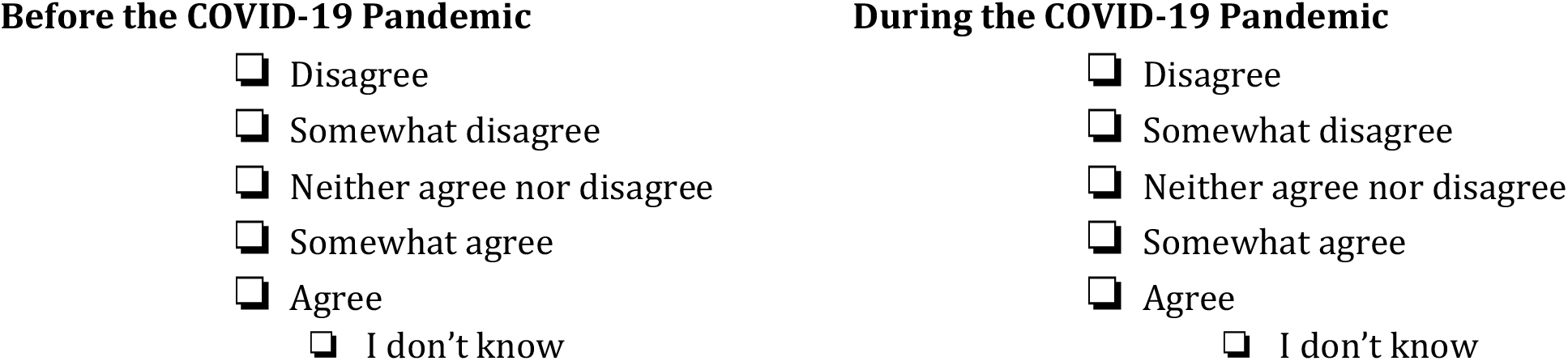

8 **The primary care team’s care coordination activities are based upon ongoing assessment of patient needs**.

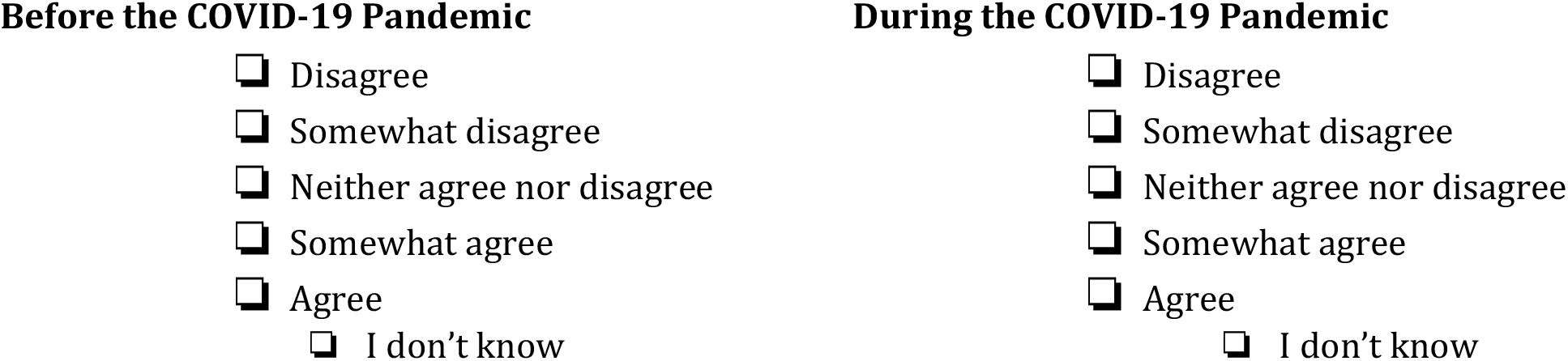

9 **The primary care team…**

a **…asks for patients’ input when making a plan for their care**.

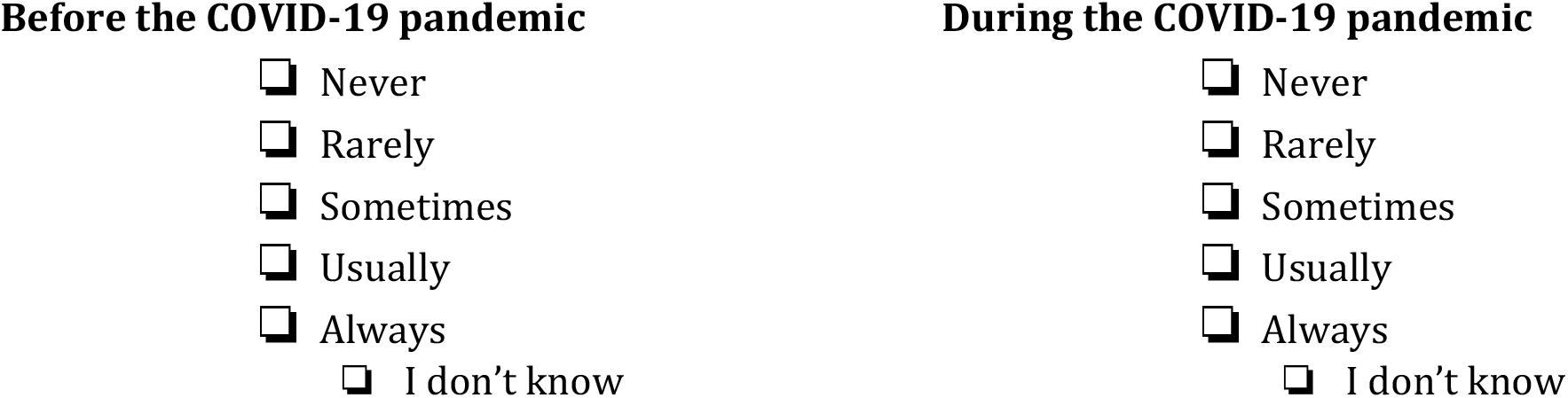

b **…helps make care plans that patients can follow in their daily life**.

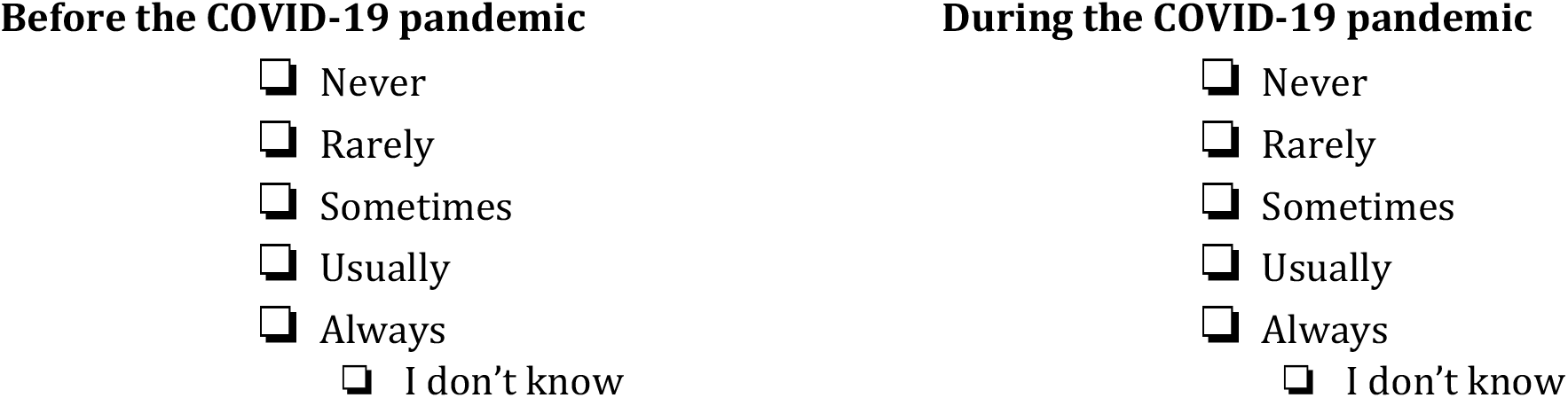

c **…develops care plans that incorporate plans recommended by other health care providers that patients see**.

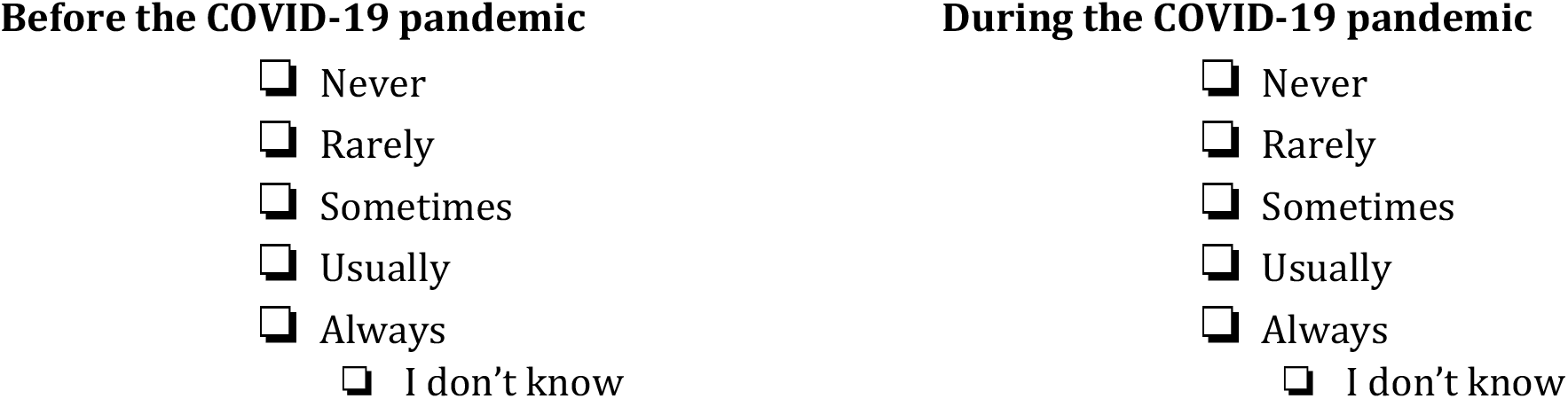

10 **The primary care team reviews and updates patients’ care plan with them**.

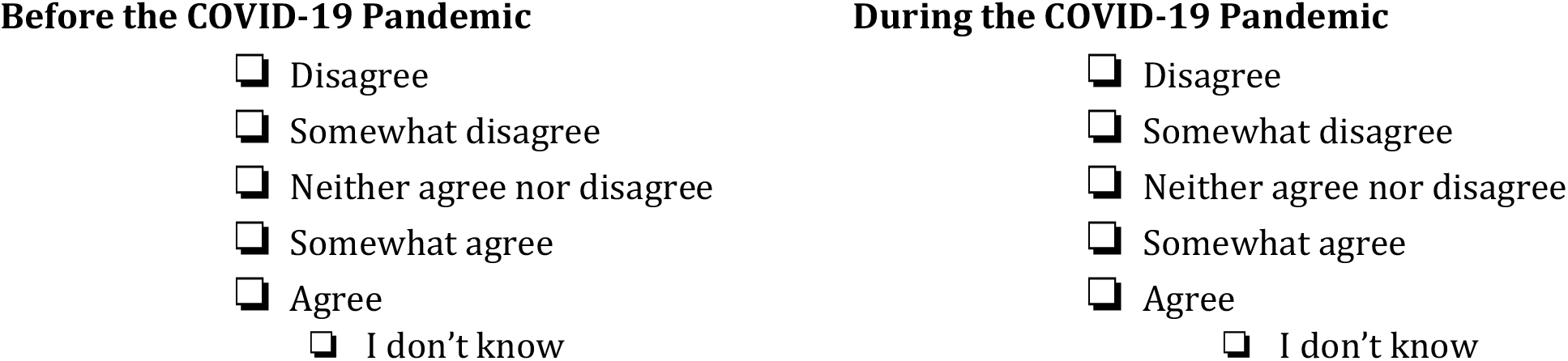

11 **The primary care team…**

a **…gives patients a copy of their care plan**.

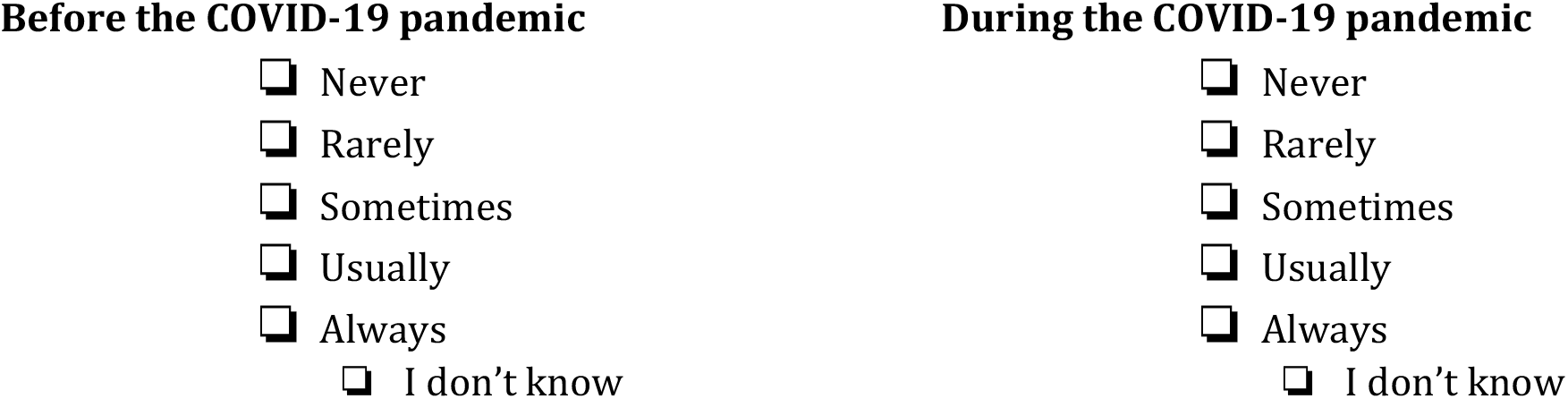

b **…follows through with the care plan**.

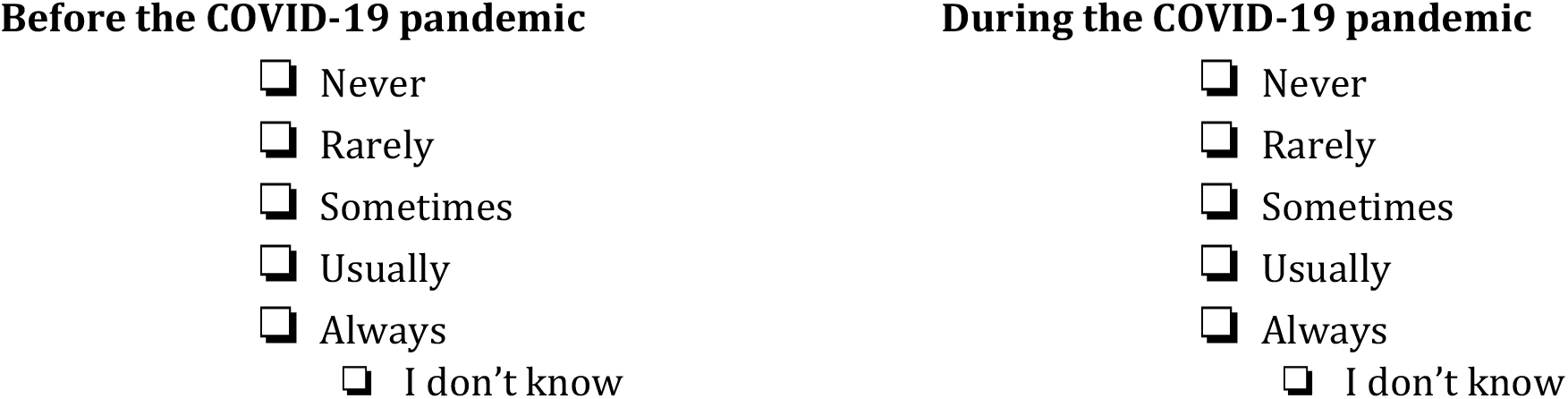

c **…uses patients’ care plan to follow progress**.

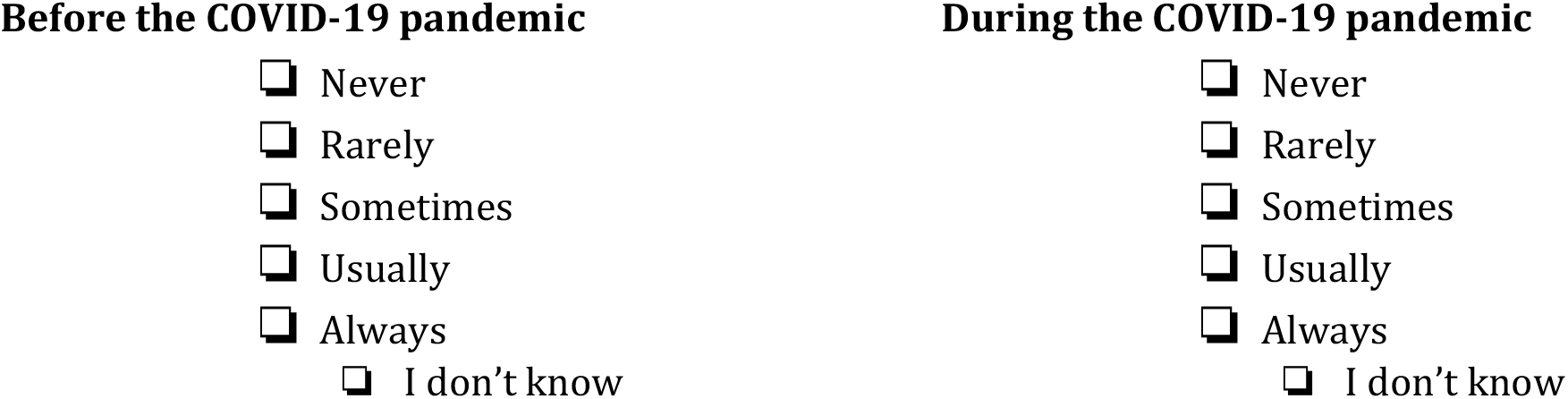

12 **The primary care team helps patients plan so they can take care of their health even when things change or when unexpected things happen**.

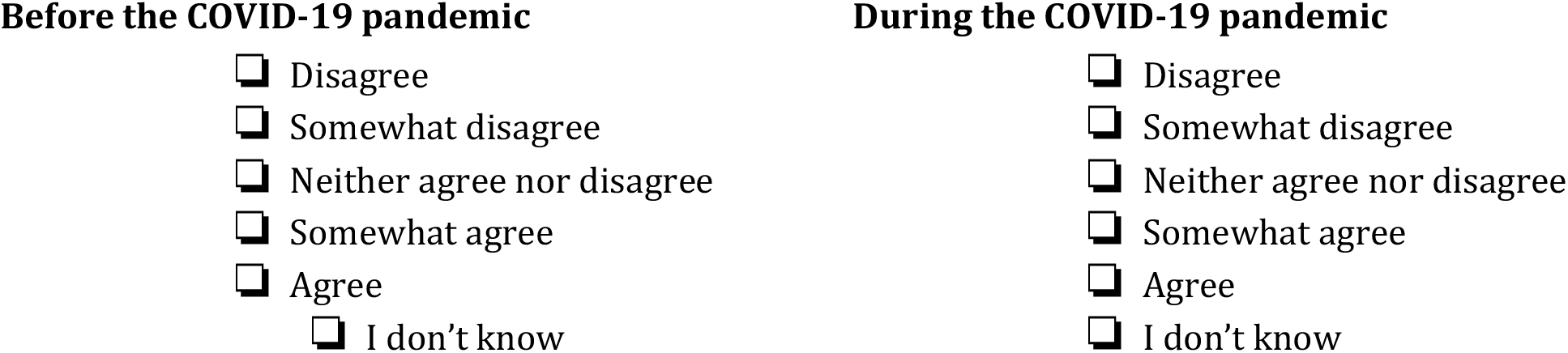

13 **Someone on the primary care team…**

a **…helps patients set goals for managing their health**.

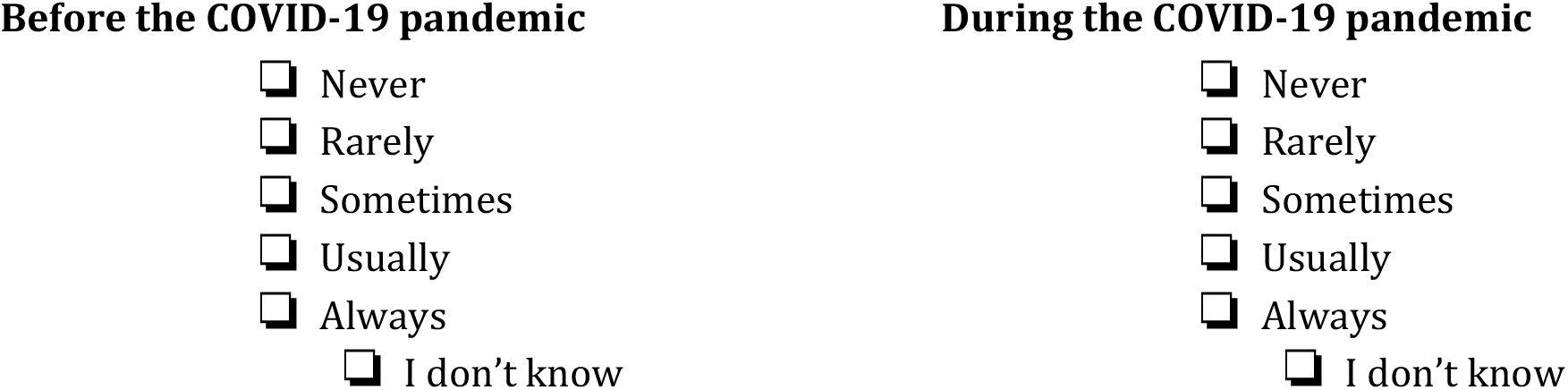

b **…checks to see if patients are reaching their goals**

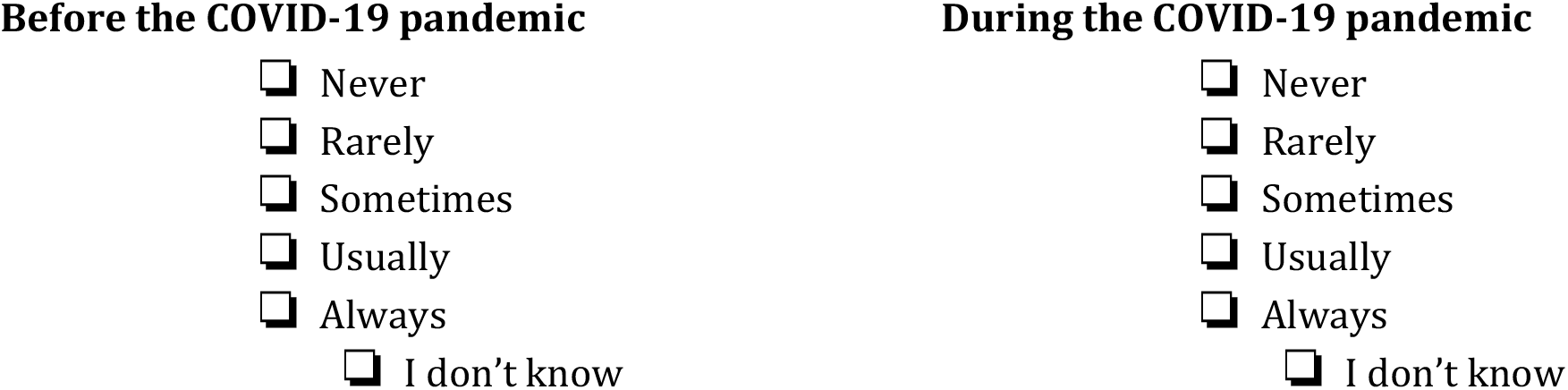

14 **The primary care practice/health centre…**

a **…has behaviour change interventions readily available for patients as part of routine care**.

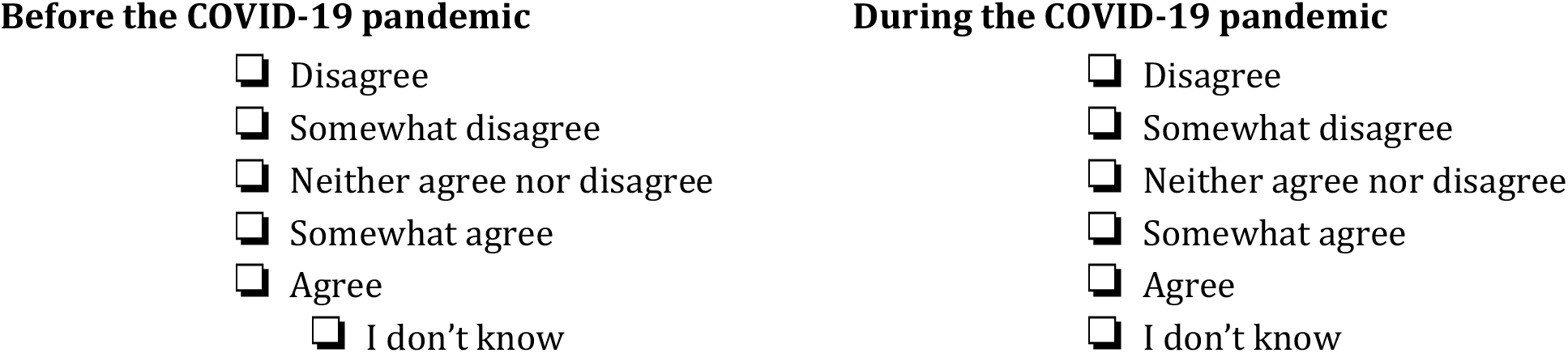

b **…has peer support readily available for patients as part of routine care**.

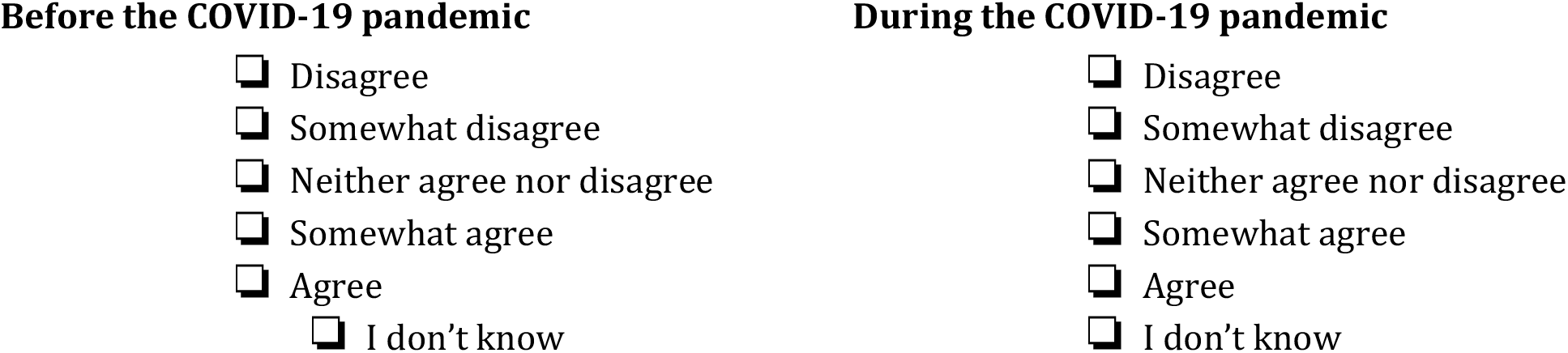

15 **Someone on the primary care team asks patients about what they need for support, such as care programs, financial services, equipment, transportation**.

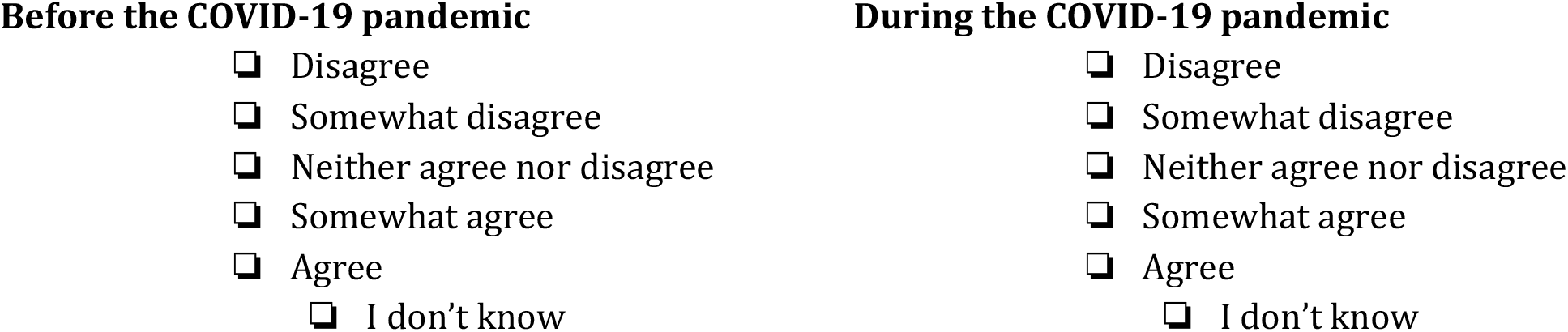

16 **Someone on the primary care team offers patients the opportunity to learn more about managing their health, such as with group appointments, support groups and patient education**.

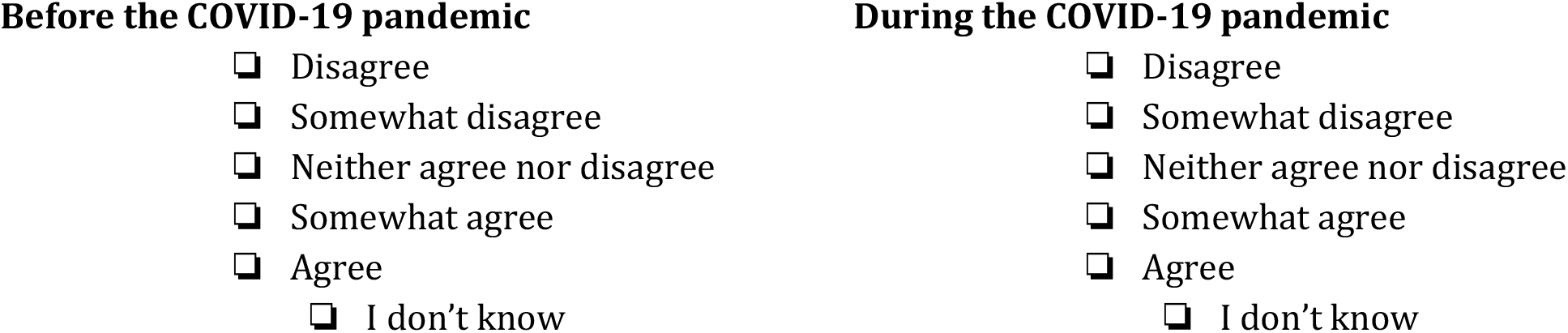

17 **Someone on the primary care team…**

a **…gives patients information about additional supportive services offered at the practice/health centre or in their community, such as counseling programs, support groups or rehabilitation programs**.

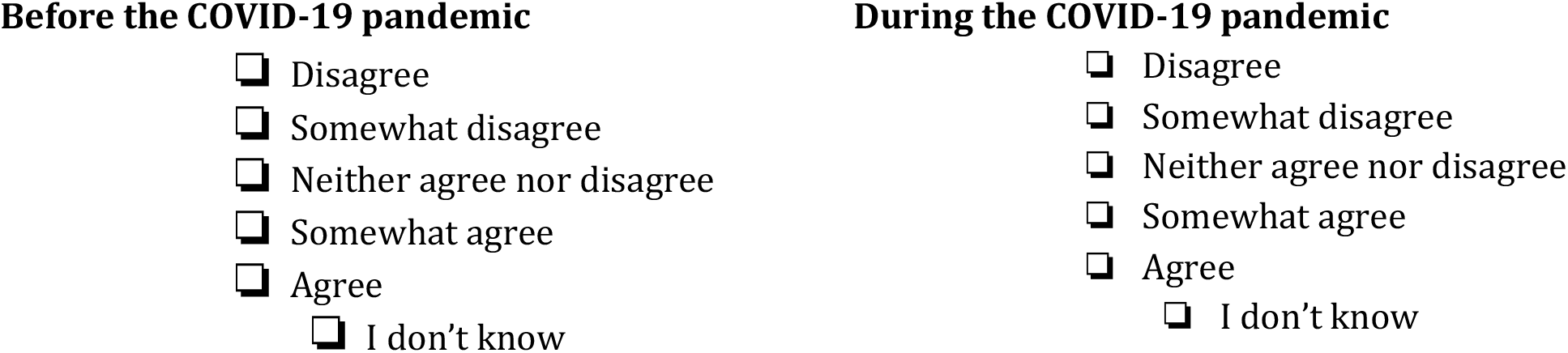

b **…encourages patients to attend programs in their community that could help them, such as support groups or exercise classes**.

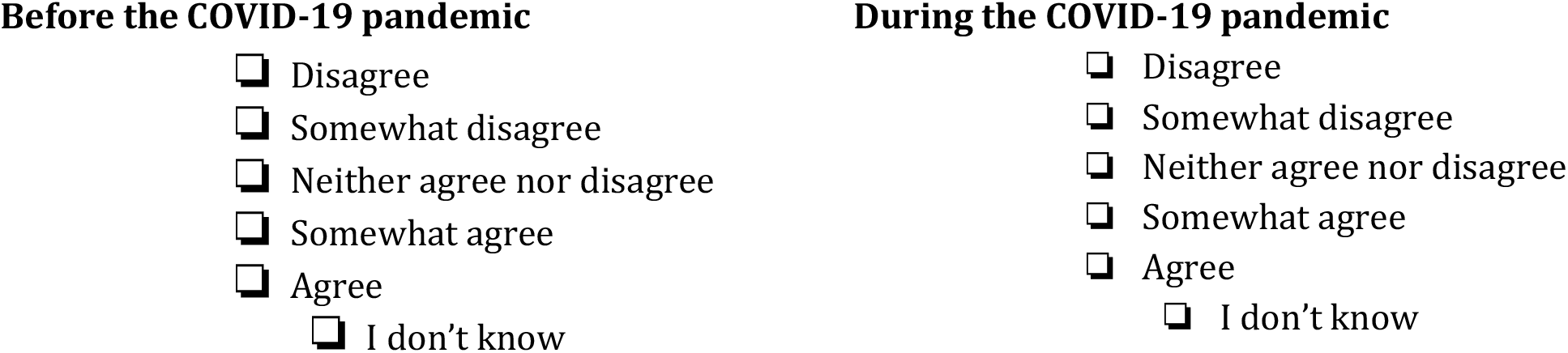

c **…connects patients to needed services, such as transportation or home care**.

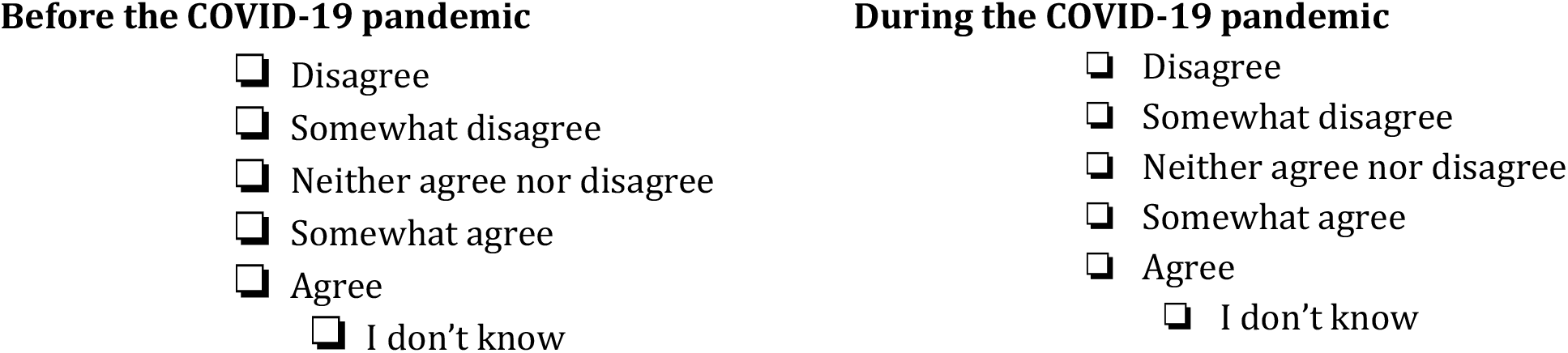

18 **When patients are discharged from the hospital, the primary care team…**

a **…is informed about the care patients received from the hospital**.

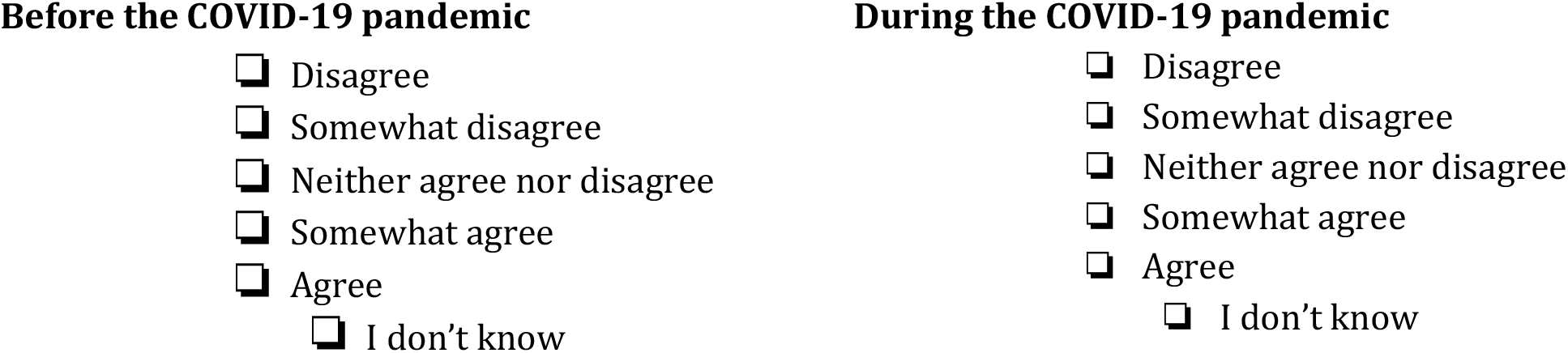

b **…receives information from the hospital about new prescriptions or if there was a change in medication**.

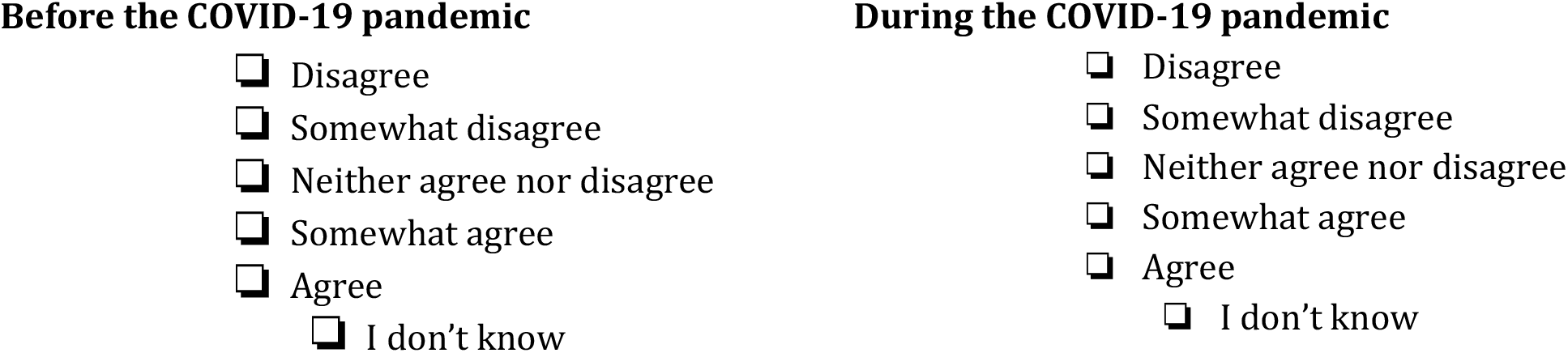

19 **When patients are discharged from the hospital, their primary care medical record includes a discharge summary in a timely manner**.

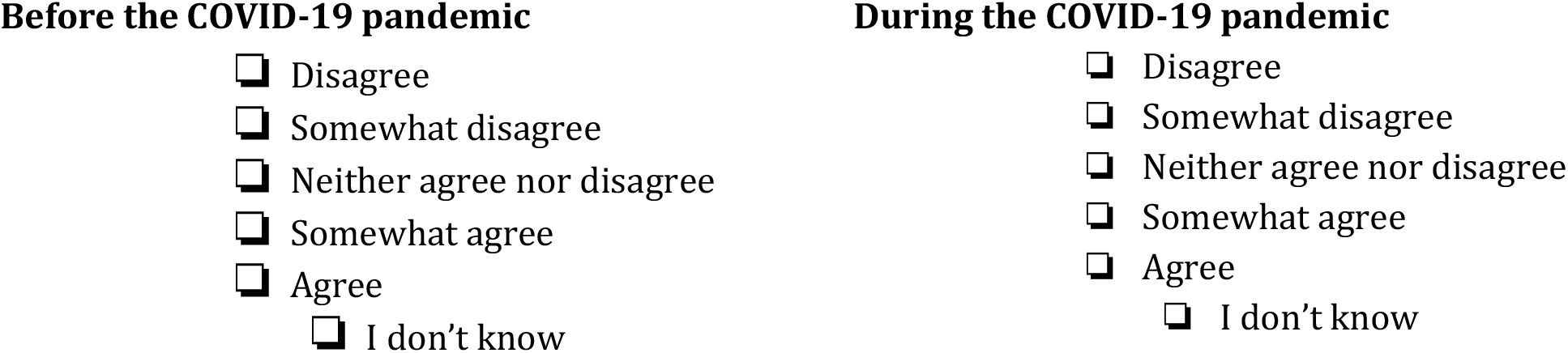

20 **When patients are discharged from the hospital and there are test results pending, their primary care medical record includes the test results within 2 weeks**.

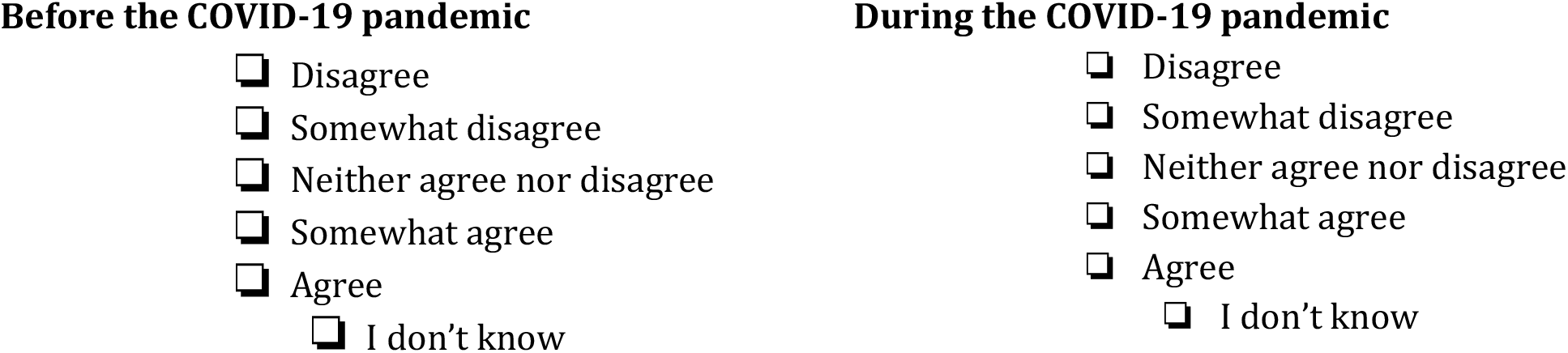

21 **In general, how would you rate the coordination of care provided at your primary care practice/health centre?**

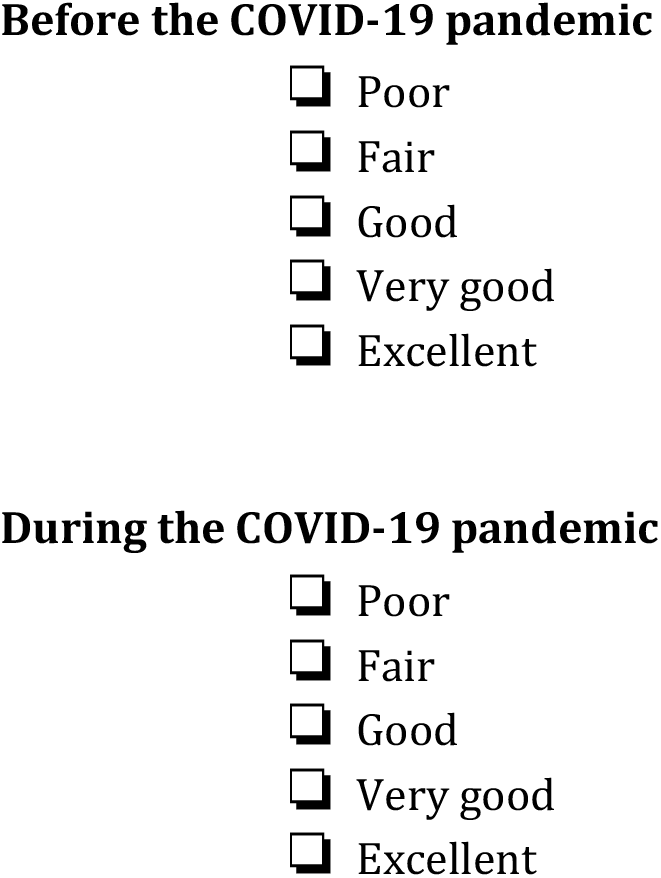

22 **What is your principal role in your health centre?** ^*^Please select all that apply
  □ Administrator
  □ Primary Care Physician
  □ Nurse Practitioner
  □ Registered Nurse
  □ Nurse Care Coordinator
  □ Other, please specify:

**Questions about your organization:**

23 **Type of Organization:**
  □ Family Health Team (FHT)
  □ Community Health Centre (CHC)
  □ Aboriginal Health Access Centre (AHC)
  □ Community Family Health Team (CHC)
  □ Nurse Practitioner Led Clinics (NPLC)

24 **Length of operation (months): _________**
25 **Number of satellite FHT sites: _________**
26 **Number of rostered patients: _________**
27 **Number of family doctors or nurse practitioners: _________**
28 **Number of Allied Health Professionals: _________**
29 **Number of family medicine residents (if applicable): _________**
30 **Who is the primary person involved with care coordination/case management?**
  □ Physician
  □ Case manager
  □ Nurses
  □ Other (specify): **_________**
  □ Unsure

31 **Do you have an EMR?**
  □ Yes
  □ No
32 **Are you using the EMR to identify patients who need care coordination/high risk patients?**
  □ Yes
  □ No
33 **Care coordination tools currently being used: *(Please check all that apply)***
  □ LACE Index Scoring Tool
  □ Predictive Repetitive Admission (PRA) Tool
  □ Probability of Repeated Admission (Pra) Risk
  □ Prediction Tool
  □ PraPlus
  □ Detection of Indicators and Vulnerabilities for Emergency Room Trips (DIVERT) Scale
  □ Number of medication +comorbidities
  □ Frailty and Vulnerability Evaluation (FAVE)
  □ Other(s) - *Please specify*:
  □ None of the above

## References

1. World Health Organization. Coronavirus disease 2019, Situation Report – 51, https://www.who.int/docs/default-source/coronaviruse/situation-reports/20200311-sitrep-51-covid-19.pdf?sfvrsn=1ba62e57_10 (2020, accessed 10 October 2022).

2. World Health Organization. Weekly epidemiological update on COVID-19 – 4, https://www.who.int/publications/m/item/weekly-epidemiological-update-on-covid-19---4-may-2023 (2023, accessed 21 July 2023).

3. Guan WJ, Liang WH, Zhao Y, et al. Comorbidity and its impact on 1590 patients with COVID-19 in China: a nationwide analysis. Eur Respir J. Epub ahead of print 26 March 2020. DOI: 10.1183/13993003.00547-2020.

4. Yang J, Zheng Y, Gou X, et al. Prevalence of comorbidities and its effects in coronavirus disease 2019 patients: a systematic review and meta-analysis. Int J Infect Dis 2020; 94: 91‐95.

5. Richardson S, Hirsch JS, Narasimhan M, et al. Presenting characteristics, comorbidities, and outcomes among 5700 patients hospitalized with COVID-19 in the New York City area. JAMA 2020; 323(20): 2052–2059.

6. Feely A, Lix LM and Reimer K. Estimating multimorbidity prevalence with the Canadian Chronic Disease Surveillance Program. Health Promot Chronic Dis Prev Can 2017; 37(7): 215–222.

7. Macdonald KM, Sundaram V, Bravata DM, et al. Closing the quality gap: a critical analysis of quality improvement strategies (Vol. 7 Care Coordination). Rockville (MD): Agency for Healthcare Research and Quality (US); 2007 Jun. (Technical Reviews, No. 9.7.) 3, Definitions of Care Coordination and Related Terms, https://www.ncbi.nlm.nih.gov/books/NBK44012/ (2007, accessed 10 October 2022).

8. Mossialos EA, Osborn R and Roland M. The Commonwealth Fund, International Experts Working Group on Patients with Complex Conditions. Designing a high-performing health care system for patients with complex needs: ten recommendations for policymakers. https://www.commonwealthfund.org/publications/fund-reports/2017/sep/designing-high-performing-health-care-system-patients-complex (2017, accessed 1 September 2022).

9. Schoen C, Osborn R, Squires D, et al. New 2011 survey of patients with complex care needs in eleven countries finds that care is often poorly coordinated. Health Aff (Millwood) 2011; 30(12): 2437–2448.

10. Lawand C, Paltser G, Cheung G, et al. Care for patients with complex needs: Canadian results from the Commonwealth Fund 2015 International Policy Survey of Primary Care Physicians. Healthc Q 2016; 19(2): 10–12.

11. Canadian Institute for Health Information. How Canada compares: results from the Commonwealth Fund’s 2019 International Health Policy Survey of Primary Care Physicians, https://www.cihi.ca/sites/default/files/document/cmwf-2019-accessible-report-en-web.pdf (2020, accessed 10 October 2022).

12. Easley J. Coordination of cancer care between family physicians and cancer specialists. Can Fam Physician 2016; 62(10): e608–e615.

13. Wang MC, Mosen D, Shuster E, et al. Association of patient-reported care coordination with patient satisfaction. J Ambul Care Manage 2015; 38(1): 69–76.

14. Kringos DS, Boerma WGW, Hutchinson A, et al. The breadth of primary care: A systematic literature review of its core dimensions. BMC Health Serv Res 2010; 10: 65.

15. Susan KM, Wallace E, O’Dowd T, et al. Interventions for improving outcomes in patients with multimorbidity in primary care and community settings. Sys Rev 2021; 10(1): 271.

16. Carroll JC, Talbot Y, Permaul JA, et al. Academic family health teams part 1: patient perceptions of core primary care domains. Can Fam Physician 2016; 62(1): e23–30.

17. Gaucher L, Dupont C, Gautier S, et al. The challenge of care coordination by midwives during the COVID-19 pandemic: a national descriptive survey. BMC Pregnancy Childbirth 2022; 22: 437.

18. Whitebird RR, Solberg LI, Jaka MM, et al. The impact of COVID-19 on patients receiving care coordination in primary care: a qualitative study. J Am Board Fam Med 2023; 36(4): 662–669.

19. Zlateva I, Anderson D, Coman E, et al. Development and validation of the Medical Home Care Coordination Survey for assessing care coordination in the primary care setting from the patient and provider perspectives. BMC Health Serv Res 2015; 15: 226.

20. Marcoux V, Chouinard M-C, Diadiou F, et al. Screening tools to identify patients with complex health needs at risk of high use of health care services: a scoping review. PLoS One 2017; 12(11): e0188663.

21. Hossain SN, Jaglal SB, Shepherd J, et al. Web-based peer support interventions for adults living with chronic conditions: scoping review. JMIR Rehabil Assist Technol 2021; 8(2): e14321.

22. Fisher EB, Ballesteros J, Bhushan N, et al. Key features of peer support in chronic disease prevention and management. Health Aff (Millwood) 2015; 34(9): 1523–1530.

23. Mangin D, Premji K, Bayoumi I, et al. Brief on Primary Care Part 2: Factors affecting primary care capacity in Ontario for pandemic response and recovery. Science Briefs of the Ontario COVID-19 Science Advisory Table 2022; 3(68).

24. Wu M-J, Zhao K and Fils-Aime F. Response rates of online surveys in published research: a meta-analysis. Comput Hum Behav Rep 2022; 7: 100206.

25. de Koning R, Egiz A, Kotecha J, et al. Survey fatigue during the COVID-19 pandemic: an analysis of neurosurgery survey response rates. Front Surg 2021; 8: 690680.

